# Prenatal smoking, alcohol and caffeine exposure and offspring externalising disorders: A systematic review and meta-analysis

**DOI:** 10.1101/2021.06.02.21258212

**Authors:** Elis Haan, Kirsten E. Westmoreland, Laura Schellhas, Hannah M. Sallis, Gemma Taylor, Luisa Zuccolo, Marcus R. Munafò

## Abstract

**Background and aims:** Several studies have indicated that there is an association between maternal prenatal substance use and offspring externalising disorders. However, it is uncertain whether this relationship is causal. Therefore, we updated a previously conducted systematic review to determine if the literature supports 1) a causal role of maternal prenatal substance use on offspring externalising disorders and 2) whether these associations differ across externalising disorders.

**Methods:** We searched Web of Science, Embase, PsycINFO and Medline databases. We included studies that examined smoking, alcohol or caffeine use during pregnancy as an exposure, and diagnosis of attention-deficit hyperactivity disorder (ADHD), conduct disorder (CD) and oppositional-defiant disorder (ODD) in offspring as an outcome. Studies on non-English language, fetal alcohol syndrome and comorbid autism spectrum disorders were excluded. Risk of bias assessment was conducted using Newcastle Ottawa Scale (NOS) and where possible meta-analysis was conducted for studies classed as low risk of bias.

**Results:** We included 63 studies. All studies were narratively synthesised, and 7 studies were meta-analysed on smoking and ADHD. The majority of studies (46 studies) investigated the association between smoking and ADHD. Studies which accounted for genetic effects indicate that the association between smoking and ADHD is unlikely to be causal. Studies on alcohol exposure in all the outcomes reported inconsistent findings and no strong conclusions on causality can be made. Studies on caffeine exposure were mostly limited to ADHD and these studies do not support a causal effect.

**Conclusions:** There is no causal relationship between maternal smoking during pregnancy and attention-deficit hyperactivity (ADHD) in offspring. However, given that the majority of identified studies investigated the association between ADHD and smoking exposure, findings with alcohol and caffeine exposures and conduct disorder (CD) and oppositional-defiant disorder (ODD) need more research, especially using more genetically sensitive designs.

## INTRODUCTION

Several studies have indicated that maternal health behaviours during pregnancy, including smoking, alcohol and caffeine consumption, may contribute to offspring externalising problems (such as attention-deficit-hyperactivity disorder (ADHD), conduct disorder (CD) and oppositional-defiant disorder (ODD)) (1-3). However, it remains unclear whether this reflects a true causal effect or residual confounding due to factors such as socioeconomic position, education, income and maternal age (4-9). This is of considerable public health importance, since smoking, alcohol and caffeine consumption are common exposures and although current UK guidelines recommend abstaining from smoking and alcohol consumption (10, 11) and limiting daily caffeine consumption to 200mg during pregnancy (12), most women still use these substances at some point in pregnancy (13, 14).

A recent systematic review (15) found evidence of an association between low to moderate maternal prenatal alcohol use and offspring behavioural and conduct problems. Similarly, systematic reviews and meta-analyses report an association between maternal prenatal smoking and offspring CD and ADHD (16-18). However, these reviews are based on conventional observational studies which do not provide strong evidence of causality, given limitations such as unmeasured and residual confounding. The only review to date to triangulate evidence from different study designs many of which are robust to confounding, concluded there was no strong evidence for an effect of maternal prenatal alcohol use on behavioural phenotypes including ADHD (19). In addition to socioeconomic confounding, the observed associations could also be explained by shared genetic influences (i.e., genetic confounding). Several studies report shared genetic liability between ADHD, CD and substance use (20-22), and maternal genetic risk for ADHD has been associated with smoking during pregnancy (23). Therefore, it is possible that the association between maternal prenatal substance use and offspring externalising disorders could be explained by genetic transmission. These reviews highlight the need for further investigation into the effect of maternal prenatal substance use on offspring externalising problems, and the use of genetically informative study designs to disentangle potential causal effects.

We systematically reviewed the evidence for association between prenatal smoking, alcohol and caffeine exposure and diagnosis of ADHD, CD and ODD. With our aim being to better understand possible causal pathways, we specifically included studies accounting for genetic effects, in addition to conventional approaches. We chose to include these three highly comorbid (24) externalising disorders to allow the interrogation of both common and specific effects of prenatal substance exposures, and prioritised clinical diagnoses over symptoms scales to avoid reporting bias.

## METHODS

We followed PRISMA and MOOSE guidelines and registered the study protocol on the Open Science Framework (10.17605/OSF.IO/D9WZK) and PROSPERO (ID: CRD42018094810).

### Search Strategy

We searched Web of Science, Embase, PsycINFO and Medline databases via the Ovid platform up to 26.04.2021 using keyword and MeSH terms (search strategy is shown in Supplementary methods), and additionally checked the reference lists of previous reviews.

### Inclusion and exclusion criteria

Inclusion criteria were: 1) publication in a peer reviewed journal in English language, 2) observational studies (cross-sectional, case-control, longitudinal, and cohort studies which also included negative control studies), 3) maternal smoking, alcohol and caffeine use measured during pregnancy, and 4) diagnosis of ADHD, CD and ODD in offspring.

Exclusion criteria for the study were: 1) animal studies, 2) reviews, 3) conference and/or meeting abstracts, 4) studies with no comparison group, 5) fetal alcohol spectrum disorder (FASD) studies, as several studies have shown an association between heavier drinking and FASD; and 6) studies with comorbid autism spectrum and tic disorders due to the different aetiology of coexistence of these disorders.

### Study selection and data extraction

Selection of studies was carried out in three stages: 1) title and abstract screening, 2) full text screening, 3) data extraction and risk of bias assessment. Study selection and data extraction took place by three reviewers: EH (100%), KW (85%) and LS (15%). Any disagreements were resolved by a third author (GT).

If studies measured multiple exposures and outcomes, data were extracted separately for each exposure and outcome. If more than one follow-up period was reported, data from the latest follow-up period were extracted (25).

### Risk of bias assessment

Risk of bias was assessed using the Newcastle Ottawa Scale (NOS) for cohorts and case-control studies (26). Studies were evaluated on three categories: 1) Selection, 2) Comparability, and 3) Outcome. Studies were ranked as low, medium or high risk of bias based on a rating system (maximum of 9 points, see Supplementary methods for details). Risk of bias assessment was conducted by the review team and points given to each study are shown in Supplementary Tables S1 and S2.

### Meta-analysis

We used random-effects models to pool results from studies at low risk of bias, and computed I^2^ statistics to quantify the between-study heterogeneity for each analysis. Statistical analyses were conducted using the metan command in Stata v.15 (27).

## RESULTS

### Literature search

After removing duplicates 5,391 articles were identified of which 393 were included in full text screening. Of these, 331 were excluded mainly due to non-specific outcome measures (n=38), wrong exposure (n=36) or not meeting outcome criteria (n=125). Excluded studies are listed in Supplementary Table S3. Study authors were contacted if relevant details were missing. Studies excluded due to missing data are listed in Supplementary Table S4. In total 63 articles were included in the current review (Figure 1).

**Figure 1.**
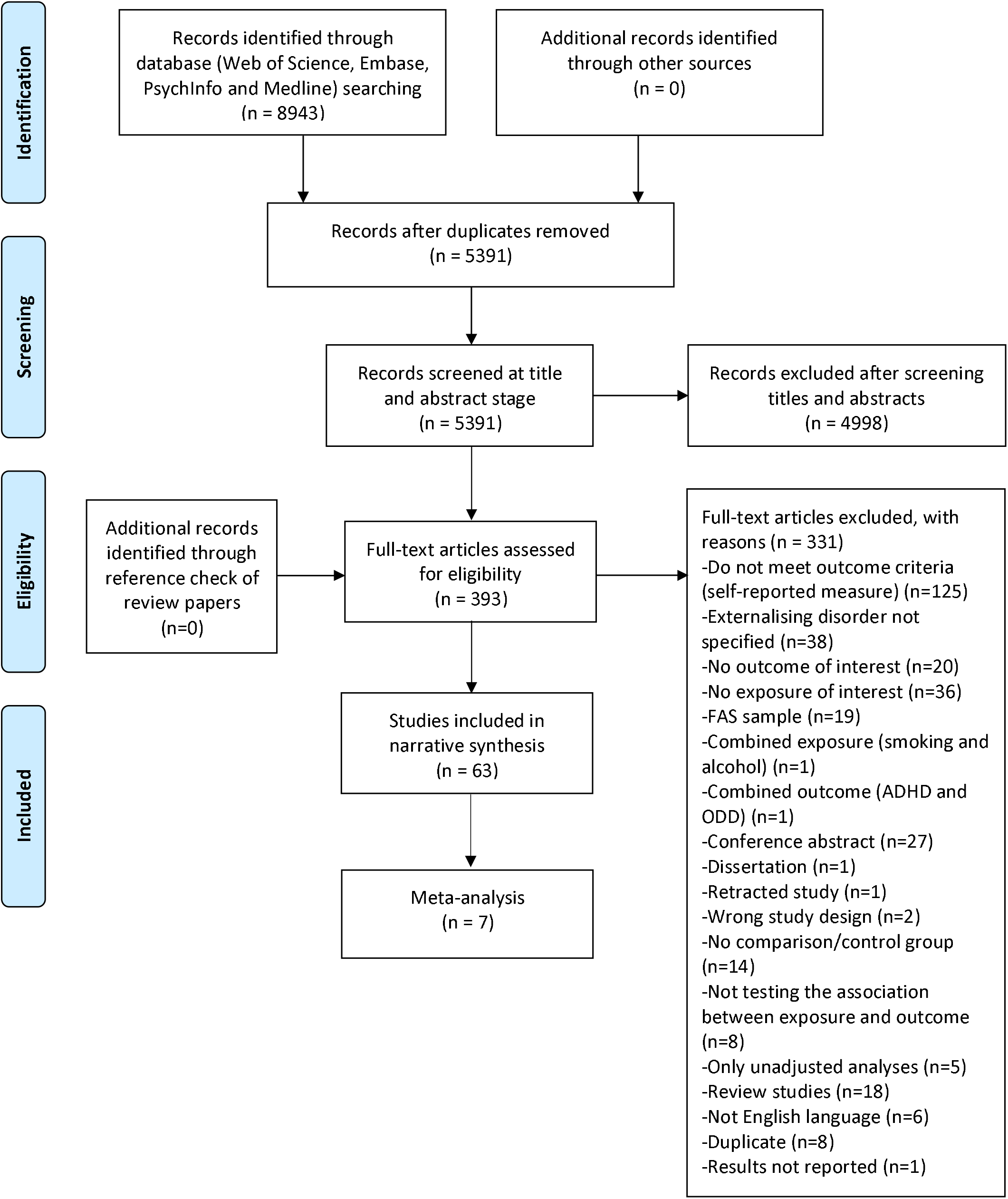
PRISMA Flowchart of search strategy.

### Characteristics of included studies

Included studies varied in terms of geographical region and study designs. The majority of studies assessed exposures retrospectively with variation in how exposures were categorised (binary vs. categorical measures). Studies also differed in follow-up time (from 2 to 37 years) and age at diagnosis (4 to 37 years). Narrative overview and full details of included studies are presented in Supplementary methods and Supplementary Tables S5 and S6.

### Inclusion of confounding variables

The majority of studies adjusted for socio-economic variables (social class, education, income, marital status), as well as for maternal age, offspring age and gender. Only a few studies adjusted for maternal mental health during pregnancy and none adjusted for partner’s substance use (see Supplementary Tables S7 and S8).

### Summary of findings

An overview of results as reported in the studies is shown in Supplementary Tables S5 and S6.

#### Prenatal smoking and ADHD

Of the 63 studies, 46 assessed the association between maternal prenatal smoking and offspring ADHD, of which 19 (41%) were cohort and longitudinal studies, 4 (9%) cross-sectional and 23 (50%) case-control studies.

Of the included cohort and longitudinal studies, 13 (68%) found a positive association between maternal prenatal smoking and offspring ADHD. Three of these studies that did not report a positive association used a sample measured when offspring were in late adolescence (16-18 years) or adulthood (37 years) (28-30).

Seven studies which found a positive association used samples from prospective longitudinal cohorts and large registries (N=5,758 to 986,046) (31-37). These studies enabled the authors to take into account environmental and/or genetic confounding by using quasi-experimental designs (i.e parental and sibling comparison designs). Six of these concluded that the association is most likely explained by confounding (31-35, 37). Two studies claimed that the association was stronger with maternal smoking compared to paternal smoking indicating a potential causal intrauterine effect (36, 38).

Seven other studies that observed a positive association adjusted for birth weight or other perinatal factors that could be potential mediators or lead to spurious association because of collider bias (39-44). Additionally, two twin studies concluded that prenatal smoking was a common risk factor among monozygotic twins concordant for ADHD (45, 46).

All four cross-sectional studies and 20 (87%) of the case-control studies found a positive association between maternal prenatal smoking and offspring ADHD. Of the three case-control studies that did not (47-49), two of these were conducted in small samples (N=372-450) (47, 48). Eight studies that observed a positive association adjusted for parental ADHD to account for potential genetic liability, but the association remained (50-57). However, another case-control study found that maternal prenatal smoking was shared between affected and unaffected siblings indicating that prenatal smoking is a weak risk factor for ADHD (58).

Seven studies examined the association with ADHD subtypes (48, 49, 51, 55, 59, 60). One study conducted in girls only found an association with hyperactive-impulsive symptoms but not inattention symptoms (51). In contrast, one study observed an indirect effect of prenatal smoking on inattention symptoms via memory span deficits (49). Two other studies focused on the inattention subtype and used the same sample (48, 55), however only one study found evidence for an association (55). Two of the remaining studies investigated gene-environment interactions in the same sample of twins and found a positive effect between maternal prenatal smoking and child genotype in children with the combined ADHD subtype (59, 60). Similarly, one study conducted in Chinese singletons found an interaction effect with all the ADHD subtypes (61). However, two other studies that also investigated gene-environment interaction – but focused on overall ADHD and used a sample of singletons – did not find an interaction effect (56, 62).

Of the six studies which investigated gender differences (29, 33, 34, 39, 63, 64), only two found evidence of a gender difference, however one study found a stronger association among girls (39), while the other found a stronger association among boys (63). Dose-response relationships were examined in 15 studies (33%), of which 12 studies observed a dose-dependent association (33-35, 37, 40, 41, 43, 55, 65-68).

##### Strength of evidence based on NOS score

In total eight longitudinal and cohort studies were rated as low risk of bias (7-9 points). Six of these were based on quasi-experimental designs (31, 33-37), one study used a twin sample (45) and another was based on a prospective cohort (69). Seven studies concluded that the association between maternal prenatal smoking and offspring ADHD is unlikely to be a causal. This was in contrast with three studies rated as very high risk of bias (0-3 points) (38, 40, 70) and three other studies rated as high risk of bias (4-5 points) (all cross-sectional designs) (39, 42, 44) which found a positive association.

In total nine case-control studies were rated as low risk of bias and eight of these studies found a positive association (47, 55-57, 64, 65, 68, 71, 72), but these studies do not account for genetic effects and can be prone to recall bias, therefore conclusions about causality should be interpreted with caution.

##### Meta-analysis based on NOS score

We conducted a meta-analysis for studies which rated as low risk of bias (7-9 points). The pooled estimate of negative control studies in maternal prenatal smoking was 1.64 (1.33-2.02) and paternal smoking 1.28 (1.19-1.39), but between-studies heterogeneity was high I^2^=79.8%. The pooled estimate of sibling comparison studies in the full sample was OR_1-9cigarettes_=1.70 (1.52-1.91); OR_>10cigarettes_=2.20 (1.78-2.73) and in the sibling matched sample OR_1-9cigarettes_=0.90 (0.83-1.11); OR_>10cigarettes_=1.04 (0.79-1.38). The pooled estimate of nested case-control studies was OR=1.61 (1.45-1.78). Results are presented in Figures 2-4.

**Figure 2.**
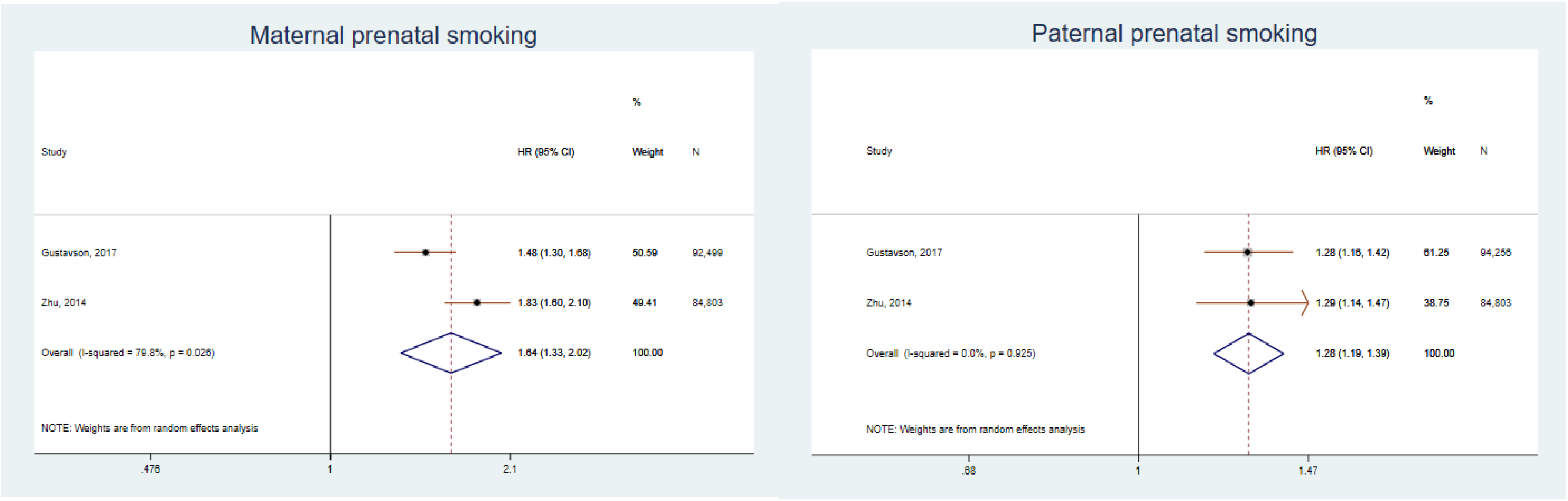
Pooled hazard ratios and 95% confidence intervals for the association between maternal and paternal prenatal smoking and ADHD. *Note: HR – Hazard ratio, 95% CI – 95% confidence intervals*

**Figure 3.**
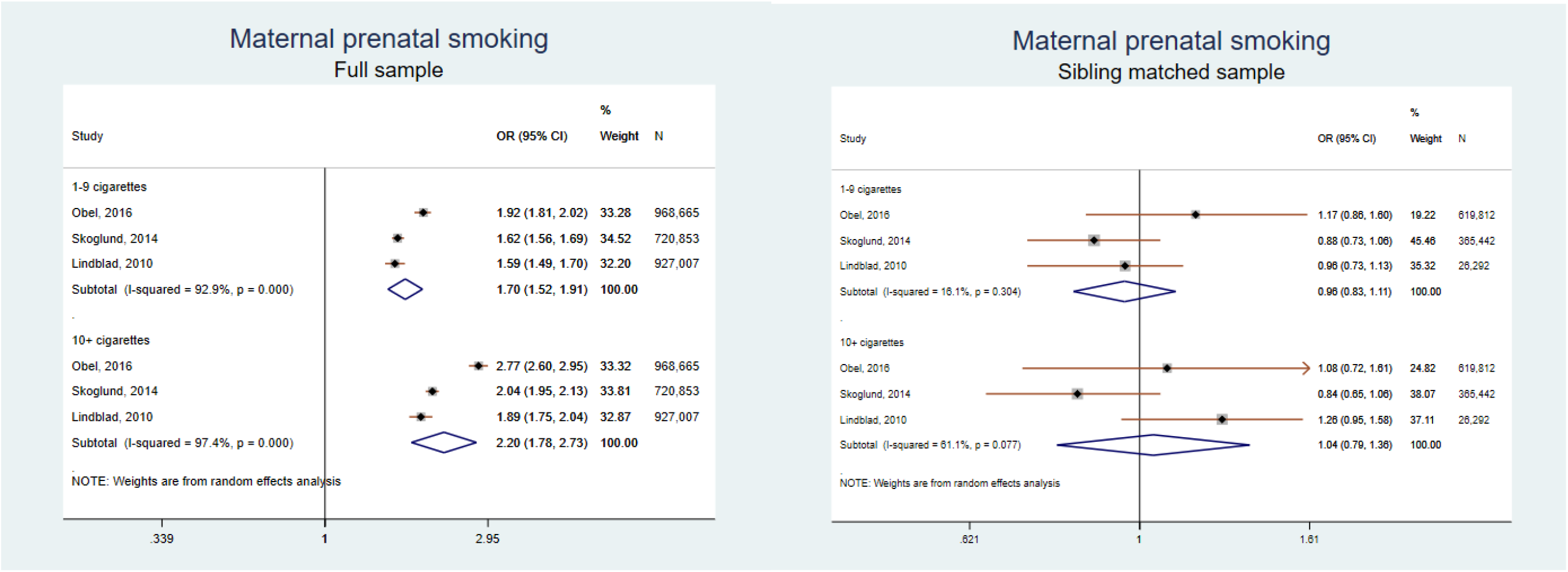
Pooled odds ratios and 95% confidence intervals for the association between full and sibling matched sample and ADHD. *Note: Studies by Obel, 2016 and Skoglund, 2014 reported results in Hazard ratios and study by Lindblad, 2010 in odds ratios (OR); 95% CI – 95% confidence intervals*

**Figure 4.**
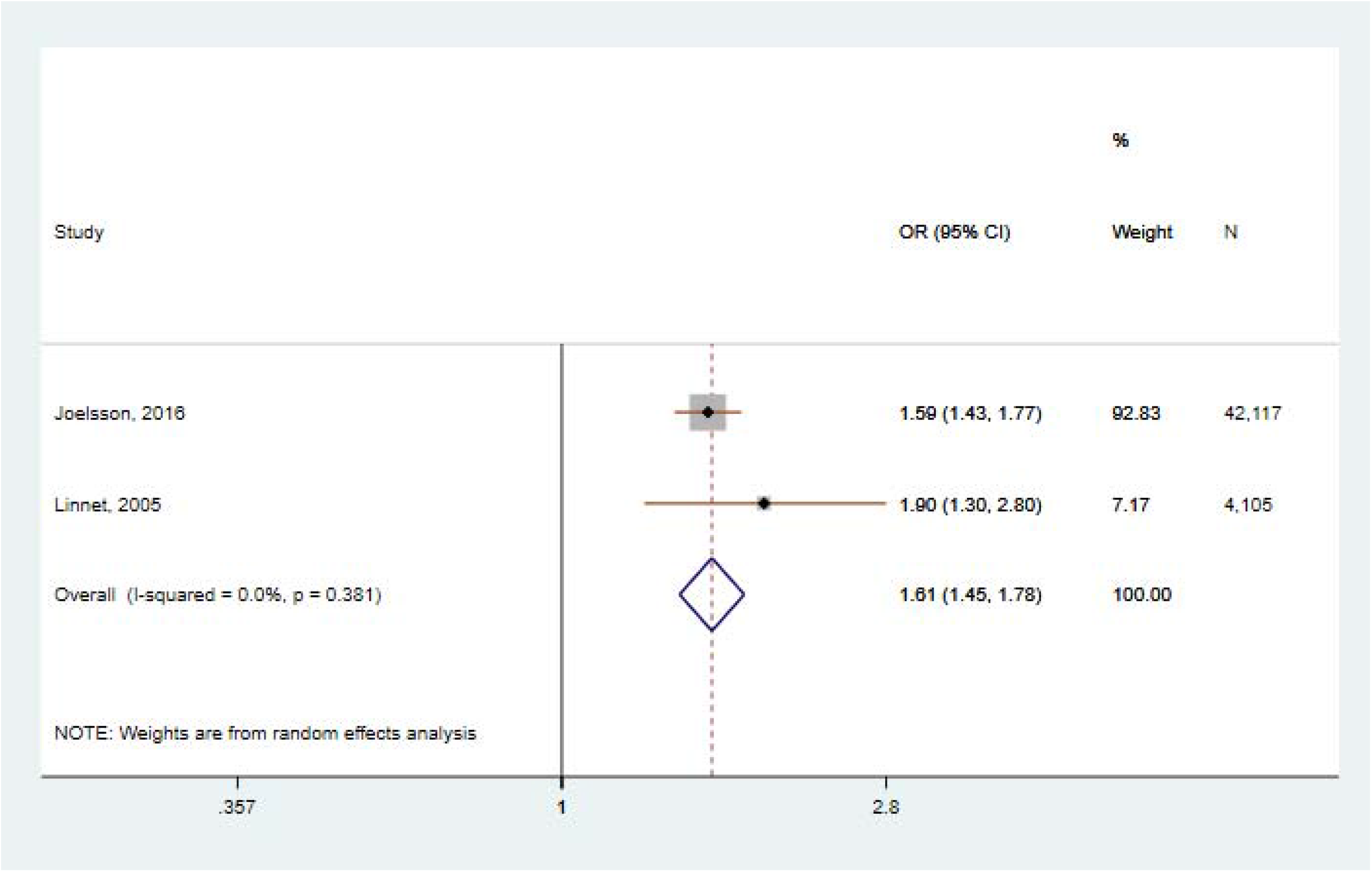
Pooled odds ratios and 95% confidence intervals for the association between nested case-control studies. *Note: Study by Joelsson reported odds ratios and study by Linnet risk ratios; 95% CI – 95% confidence intervals*

#### Prenatal smoking and CD and ODD

10 studies investigated the association between maternal prenatal smoking and offspring CD, of which 5 were cohort and longitudinal studies, 2 cross-sectional studies and 3 case-control studies. Six studies (60%) found an association between maternal prenatal smoking and offspring CD (50, 73-77). However, four of these studies used a clinical or hospital referred sample (29, 50, 76, 77). One study observed an interaction effect between maternal prenatal smoking and child genotype (76). In the three studies that did not find evidence of an association, two studies used a sample of offspring in late adolescence (16-18 years) (28, 29) and one study found an indirect effect via neuropsychological functioning (49).

Six studies investigated the association between maternal prenatal smoking and offspring ODD of which three were cohort and longitudinal studies, one cross-sectional study and two were case-control studies. Two studies (N=798-995) found an association with maternal prenatal smoking (75, 78). Among the studies that did not observe an association, one found an indirect effect via neuropsychological functioning, similar to the effect observed for ADHD and CD (49). Two studies measured ODD in adolescence (15 years) and adulthood (21 years) where disorder manifestation could differ from childhood (77, 79). One other study was conducted in a small sample (N=215) and may have lacked power to detect an effect (38).

##### Strength of evidence based on NOS score

Only one study based on smoking and CD was rated as low risk of bias (8 points) and this study did not find evidence for an association between prenatal smoking and CD (28). Two studies rated as very high risk of bias (2-3 points) did not find an association between prenatal smoking and ODD (38, 79). Other studies rated as high risk of bias (4-6 points) found an association between prenatal smoking and offspring CD and ODD (29, 50, 63, 73-76, 78), but two studies were based on cross-sectional or case-control design which cannot prove causality (50, 74) and other four studies used a clinical or hospital referred sample (29, 63, 73, 76).

#### Prenatal alcohol and ADHD

Thirteen studies investigated the association between maternal prenatal alcohol consumption and offspring ADHD of which 8 were cohort, longitudinal and cross-sectional studies (N=679-34,503), 1 was longitudinal twin study (N=1,936) and 4 were case-control studies (N=372-2,419). Two longitudinal studies found a positive association only with heavier alcohol use (46, 80) and one other longitudinal study found a positive association with alcohol use in all trimesters and with binge drinking (81), but this study was conducted in a small sample (N=81). Three (33%) case-control studies found a positive association with maternal prenatal alcohol consumption (54, 66, 82) of which one used heavier drinking (drunkenness during the first 2 months) as the exposure (66). Of the two other studies, one was conducted in a hospital referred sample (54) and the other failed to adjust for many relevant confounders (82).

##### Strength of evidence based on NOS score

Two studies based on alcohol exposure and ADHD were rated as low risk of bias (8-9 points) and these did not find evidence for an association (69, 83). Four longitudinal cohort studies (41, 81, 84, 85) were rated as high risk of bias (4-6 points), however only the highest scoring study (4 points) found evidence of an association (81). Out of four case-control studies rated as high risk of bias (5-6 points), three studies reported an association between prenatal alcohol exposure and offspring ADHD (54, 66, 82). Due to the variability on exposure assessment, a meta-analysis was not possible.

#### Prenatal alcohol and CD and ODD

Five cohort, longitudinal and cross-sectional studies investigated the association between maternal prenatal alcohol consumption and offspring CD and ODD (N=546-9,719). Four studies were on CD (28, 80, 86, 87) and two on ODD (79, 80). One study observed a positive association with heavier drinking and ODD (80), and two studies found an association between maternal prenatal alcohol consumption and offspring CD (86, 87). However, these studies used heavier alcohol consumption and binge drinking phenotypes or were based on samples from culturally distinct populations (87).

##### Strength of evidence based on NOS score

Two studies based on alcohol exposure and CD were rated as low risk of bias (8 points). One of these studies did not find evidence for an association (28), but the other found evidence for an association with heavier alcohol use (86). Both of these studies were based on prospective longitudinal birth cohorts. Two studies were rated as very high risk of bias (2 points), one study on CD found a positive association with binge drinking (87) while the other investigating ODD and using cross-sectional design did not find evidence for an association (79).

#### Prenatal caffeine and ADHD and ODD

Three studies investigated the association between maternal prenatal caffeine consumption and offspring ADHD (N=3,627-24,156) (82, 88, 89). One study examined the association with offspring ODD (N=5,924) (79). No evidence for an association was observed between maternal prenatal caffeine consumption and offspring ADHD. Two of these studies used a longitudinal cohort design (88, 89) and one study used a case-control design (82). A study of ODD based on a cross-sectional sample found weak evidence for an association with maternal prenatal caffeine use in girls (79).

##### Strength of evidence based on NOS score

Two studies based on caffeine exposure and ADHD were rated as low risk of bias (8-9 points) (88, 89). These studies found no evidence for an association. One case-control study rated as high risk of bias (4 points) also did not find evidence for an association (82). Only one study on ODD rated as very high risk of bias (2 points) found weak evidence of an association in girls (79). The studies at low risk of bias assessed caffeine consumption differently (one study in mg derived from coffee and tea/mate and other study in cups of coffee) and could not be meta-analysed.

## DISCUSSION

In this systematic review we examined whether there is evidence to support a causal effect of maternal prenatal smoking, alcohol and caffeine use on offspring ADHD, CD and ODD risk by synthesising the results of existing research based on risk of bias assessment. Overall, our findings support stronger associations between prenatal smoking and ADHD and CD. However, evidence was less clear for the association with ODD and inconsistent on alcohol exposure for all outcomes. Our findings on caffeine exposure were limited to ADHD and there was lack of evidence for other outcomes.

Our findings for smoking exposure indicate that maternal prenatal smoking is more strongly associated with ADHD and CD than with ODD. However, given that there were few studies on ODD, no strong conclusions can be drawn. Furthermore, some studies on ADHD with low risk of bias were able to take into account genetic effects, and indicate that shared genetics plays a substantial role in the association with prenatal smoking. This is supported by a previous systematic review based on genetically informed designs which also concluded that the association between maternal prenatal smoking and ADHD and CD symptoms is explained by familial confounding and shared genetics (90).

We identified relatively few studies that investigated the association between prenatal alcohol exposure and diagnosis of ADHD, CD and ODD in offspring. Evidence from these studies indicates that an association exists between heavier alcohol consumption and ADHD and CD. A recent review and meta-analysis on low to moderate maternal alcohol consumption during pregnancy and offspring ADHD did not find evidence for an increased risk of ADHD symptoms (91), but studies on CD symptoms using quasi-experimental designs have found evidence for a potential causal effect (92, 93). However, these studies may be biased since outcome measures are maternally reported (94). Similarly to alcohol exposure, we only identified a few studies on prenatal caffeine exposure and these studies do not provide evidence for a causal effect with ADHD.

Several weaknesses and sources of heterogeneity between included studies emerged while we appraised the current research. First, studies varied greatly on number of confounders adjusted in the multivariable analyses, thus raising the possibility of residual confounding. Although many studies adjusted for socioeconomic factors known to affect both exposures and outcomes, none of the studies adjusted for partner’s substance use during pregnancy. There is evidence that assortative mating affects parental smoking and alcohol consumption and failure to take into account partner’s substance use can lead to biased effect estimates (95). Similarly, only a limited number of studies accounted for maternal mental health during pregnancy. For example, it has been shown that maternal depressive and anxiety symptoms during pregnancy increase risk of offspring behavioural problems (96). In contrast, many studies on smoking exposure adjusted for perinatal factors, such as birth weight, gestational age or other pregnancy and birth complications, which could be potential mediators in the pathway between prenatal smoking and ADHD. Adjusting for mediators induces collider bias in unpredictable directions, as showed in previous studies (97). Therefore estimates adjusted for birth weight may result in spurious association if there is an unmeasured common cause between birth weight and outcome (98).

Second, maternal prenatal exposure assessment was mostly based on self-reports and mothers may under report their prenatal substance use, due to social desirability, which may lead to biased effect estimates in the studies. Furthermore, many studies assessed exposures after the child’s birth or retrospectively when the outcome was already present, which may lead to recall bias (this is the case for all included cross-sectional and for most case-control studies). In these studies, causality should be interpreted cautiously. Third, studies also differed in terms of how prenatal smoking, alcohol and caffeine consumption were categorised. Many studies used a binary measure which does not adequately capture effects of substance use where these are dose dependent. Some studies used a scale of low, moderate and high, but there is no clear definition of the level of consumption each of these categories represent. Fourth, studies also varied on timing of substance use with the majority of studies using a single time point assuming that the effects of maternal substance use remain constant throughout pregnancy. One study on alcohol exposure reported that maternal prenatal alcohol consumption had a more harmful effect on offspring CD during the 1^st^ trimester compared to the 3^rd^ trimester indicating that prenatal alcohol exposure during the 1^st^ pregnancy trimester may be more harmful (86).

Fifth, considering that there is a high comorbidity between externalising disorders, few studies took this into account. Although high rates of comorbidity are common among psychiatric disorders, it is plausible that a somewhat different aetiology may underlie externalising disorders with and without comorbidities. For example, two studies that observed the association between maternal prenatal smoking and ADHD with comorbid conditions found that ADHD with comorbid CD/ODD had a stronger association with maternal prenatal smoking than ADHD without comorbidities (57, 71).

Sixth, although externalising disorders are more prevalent among boys than girls (99) few studies investigated gender effects. Some studies have shown that boys exposed to prenatal smoking and alcohol consumption may be at higher risk for developing behavioural problems than girls (100, 101) and it is possible that prenatal substance use may have distinct effects on boys and girls which needs more research.

Seventh, studies varied greatly on age when ADHD, CD and ODD were assessed. Although several studies have shown that childhood externalising disorder symptoms persist into adulthood (102, 103), other studies have found that childhood mental health problems change across development and persistence of externalising disorders depends on severity and comorbidity of symptoms (104, 105). Previous studies on ADHD have reported that presentation of hyperactive-impulsive and inattention symptoms varies from preschool to early adulthood and inattention symptoms tend to be more persistent (106). It is also suggested that child- and adulthood ADHD are two separate diagnoses and future studies should investigate which underlying mechanisms could explain these different developmental paths (107).

One major strength of this systematic review is including multiple prenatal exposures (smoking, alcohol and caffeine) and outcomes (ADHD, CD and ODD) which enabled us to synthesise whether the associations would differ across different exposure-outcome combinations. Additionally, conducting risk of bias assessments enabled us to account for potential weaknesses in study designs when interpreting the evidence supporting a causal relationship.

However, this systematic review also has some limitations. First, we limited the searches to studies that used diagnosis as an outcome measure, and therefore we missed studies reporting on symptoms scores or other continuous scales. Second, we only included English language studies. However, it has been shown that the exclusion of non-English studies has a little impact on overall findings (108).

### Conclusion

Our review showed that there is an association between maternal prenatal smoking and offspring ADHD, but studies that accounted for shared genetic and environmental confounders suggest that this association is unlikely to be causal. Given that majority of the identified studies investigated the association between ADHD and smoking exposure, findings with alcohol and caffeine exposures and CD and ODD need more research, especially using genetically sensitive designs. Future studies should use more prospective and quantitative exposure measures during each pregnancy trimester, as well as take into account comorbidities between externalising disorders, gender differences and changes in presentation and manifestation of externalising disorder symptoms across development.

## Data Availability

All data we used for this manuscript are available in text, tables, figures and supplementary file.

