## Supplementary material for "Prenatal smoking, alcohol and caffeine exposure and offspring externalising disorders: A systematic review and meta-analysis"

**Supplementary methods**

The search strategy:

1. Outcome terms: "attention deficit disorder" OR "hyperactiv*" OR "impulsiv*" OR "ADHD" OR "attention deficit hyperactivity" OR “ADD” OR "externali?ing" OR “conduct disorder” OR “behavio* disorders” OR “behavio* problem*” OR “disruptive behavio*” OR “oppositional defiant disorder”.
2. Exposure terms: "alcohol*" OR "tobacco" OR "caffein*" OR "drink*" OR "ethanol" OR “drinking” OR "smoking" OR "cigarette*" OR "nicotine" OR "coffee" OR "tea" OR "energy drink*"OR “taurine”
3. Population terms: “pregnan*” OR “perinatal” OR “prenatal” OR “intrauterine*” OR “utero” OR” f?etal” OR “gestation” OR “trimester”

Search performed in Medline (Ovid platform) including relevant MeSH Terms

1. disruptive, impulse control, and conduct disorders"/ or "attention deficit and disruptive behavior disorders"/ or attention deficit disorder with hyperactivity/ or conduct disorder/ or child behavior disorders/ - MeSH Terms
2. Child Behavior Disorders/ - MeSH Terms
3. Impulsive Behavior/ - MeSH Terms
4. Problem Behavior/ - MeSH Terms
5. attention deficit disorder.tw
6. hyperactiv*.tw
7. impulsiv*.tw
8. ADHD.tw
9. attention deficit hyperactivity.tw
10. externali?ing.tw
11. conduct disorder.tw
12. behavio* disorders.tw
13. behavio* problem*.tw
14. disruptive behavio*.tw
15. oppositional defiant disorder.tw
16. ALCOHOLS/ - MeSH Terms
17. DRINKING BEHAVIOR/ or ALCOHOL DRINKING/ or DRINKING/ or BINGE DRINKING/ - MeSH Terms
18. TOBACCO USE"/ or TOBACCO/ or TOBACCO SMOKING – MeSH Terms
19. CAFFEINE/ - MeSH Terms
20. COFFEE/ - MeSH Terms
21. TEA/ - MeSH Terms
22. Energy Drinks/ - MeSH Terms
23. Ethanol – MeSH Terms
24. Taurine – MeSH Terms
25. alcohol*.tw
26. tobacco.tw
27. caffein*.tw
28. drink*.tw
29. ethanol.tw
30. smoking.tw
31. cigarette*.tw
32. nicotine.tw
33. coffee.tw
34. tea.tw
35. energy.drink*.tw
36. taurine.tw
37. Pregnancy trimesters – MeSH Terms
38. Pregnancy – MeSH Terms
39. PREGNANCY TRIMESTER, SECOND/ or PREGNANCY TRIMESTER, FIRST/ or PREGNANCY TRIMESTER, THIRD/ - MeSH Terms
40. Pregnan*.tw
41. Perinatal*.tw
42. Prenatal*.tw
43. Intrauterine*.tw
44. Utero.tw
45. F?etal.tw
46. Gestation.tw
47. Trimester.tw

Narrative overview of included studies

Of the included studies, 27 were North American, 24 European of which 14 were Scandinavian, 4 Australian, 5 South American and 3 Asian. Of the included studies, 53 measured smoking exposure, 17 measured alcohol exposure and 4 studies measured caffeine exposure.

Of the included 33 cohort and longitudinal studies and 6 cross-sectional studies, 6 studies were based on selective populations, such as participants from hospital and indigenous culture, areas of lower socioeconomic status, as well as participants of depressed probands. Additionally, three studies were conducted in twins. Of the 24 case-control studies included, 6 studies selected cases and controls from clinical and/or hospital-based sample and 2 studies were conducted in twins.

Follow-up time ranged from 2 to 37 years in cohort and longitudinal studies. Out of the 63 included studies, eight studies reported results separately for boys and girls (N=147 to 968,665), two studies were conducted only in boys (N=177 to 400) and two studies only in girls (N=228 to 1,936).

Assessment of exposures

*Smoking*

Of the 53 studies on smoking exposure, 23 studies (43%) used cohort and/or longitudinal design, 6 studies (11%) used cross-sectional, and 24 studies (45%) used case-control study design (Supplementary Table S5 & S6). Sample sizes ranged from 147 to 968,665 participants. A total of nine (39%) of the included cohort studies used a binary exposure measure of which three studies used a cut-off <10 cigarettes per day. Of the cross-sectional studies, four studies used a binary exposure measure. 16 (57%) studies used different categorical measures of which one study included occasional smokers to the group of no smokers. Additionally, one case-control study used cotinine as a measure for smoking exposure.

A total of 12 (52%) of the included cohort and longitudinal studies assessed smoking during pregnancy prospectively. Three of these studies measured smoking in each pregnancy trimester, three studies in two pregnancy trimesters and the remaining three studies measured smoking at one time point during pregnancy. 12 (56%) studies assessed smoking some years after child’s birth. Four cross-sectional studies assessed smoking retrospectively and two studies did not provide these details.

Of the 24 included case-control studies, 5 studies (21%) assessed smoking prospectively and 19 (79%) studies retrospectively. Three studies (14%) assessed smoking in each pregnancy trimester, two studies during the first and/or second pregnancy trimester and 18 (82%) studies did not provide details of the pregnancy trimester during which smoking was assessed.

*Alcohol*

Of the 17 studies on alcohol exposure, 11 (65%) were cohort and longitudinal studies, 2 (12%) used cross-sectional design and 4 (23%) were case-control studies. Total sample size ranged from 81 to 48,072 participants.

Of the 13 included cohort, longitudinal and cross-sectional studies, 9 studies (69%) used different categorical measures and 3 studies (23%) used a binary measure. One longitudinal cohort study measured alcohol exposure using the continuous score from the AUDIT scale. Of the 4 case-control studies included, 3 studies used a binary measure, and one study used a categorical measure.

Out of these 17 studies, 5 studies (23%) assessed alcohol consumption prospectively, 10 studies (69%) retrospectively, one study few months after child’s birth and one other study did not provide these details. Two studies assessed alcohol consumption in each pregnancy trimester, two studies during the 1^st^ pregnancy trimester and other two during the 1^st^ and 3^rd^ pregnancy trimester or 1^st^ trimester and beyond.

*Caffeine*

Of the 4 studies on caffeine exposure, 2 used a cohort study design, 1 study was cross-sectional and the other used a case-control design. The total sample size ranged from 2,419 to 24,068 participants.

Two studies used categorical measure of caffeine exposure, of which one study included ‘some’ caffeine consumption as a baseline. Two other studies used a binary measure of which one used none to less than one cup per day as a reference. Two studies derived daily caffeine consumption from coffee and tea/mate intake, 1 study only from coffee and one study did not specify the source of caffeine in their analyses. Two studies assessed caffeine consumption prospectively of which one during the 2^nd^ pregnancy trimester. The other two studies measured caffeine consumption retrospectively, of which one study asked about caffeine consumption in each pregnancy trimester and the other study did not specify in which pregnancy trimester caffeine intake was assessed.

Assessment of outcomes

*ADHD*

Of the 63 included studies, 54 studies measured ADHD. Of these studies, two focused only on ADHD inattentive subtype and four ADHD combined or hyperactive-impulsive and inattentive subtypes.

Sixteen studies (30%) used hospital or national registry databases for ADHD diagnosis and/or ADHD medication use. 25 studies (46%) used diagnostic interviews and most commonly the Diagnostic Interview Schedule for Children (DISC) and the Schedule for Affective Disorders and Schizophrenia (K-SADS) were used. Additionally, the Missouri Assessment of Genetics Interview for Children (MAGIC) and the Preschool Age Psychiatric Assessment (PAPA) were also used. Furthermore, two studies used the Development And Well-Being Assessment (DAWBA) maternal and teacher reports for generating ADHD diagnosis called “DAWBA bands”, six studies (11%) derived ADHD diagnosis based on clinical assessment or by using multiple evaluations by mothers, teachers and clinicians. Five studies (9%) used parent report of whether their child had been diagnosed with ADHD.

Offspring age at assessment varied from 4 to 37 years. 5 studies were assessed at offspring age below 6 years, 39 studies were assessed at offspring age between 6 to 13 years, 5 studies at offspring age 14 to 18 years, 4 studies were assessed in adulthood (at age 20 to 37 years) and one study did not provide these details.

*CD and ODD*

Of the 63 studies, 13 studies measured conduct disorder and 7 studies measured ODD. All studies derived diagnosis using diagnostic interviews (either DISC, K-SADS or PAPA) and the World Health Organization’s Composite International Diagnostic Interview (CIDI).

Offspring age at CD assessment varied from 6 to 21 years. Five studies were assessed at offspring age from 8 to 15 years, six studies from 6 to 17 years, one study was assessed in adulthood and one other study did not provide these details. Offspring age at ODD assessment varied from 4 to 21 years. Two studies assessed ODD when offspring were aged 4 years, four studies at offspring age 10 to 17 years, and one study in adulthood (21 years).

Risk of bias assessment

### Newcastle Ottawa Scale for cohort studies

| **Item** | **Points** |
| --- | --- |
| **Selection** | |
| 1) Representativeness of the exposed cohort | 1 point – cohort is representative of the average pregnant woman in the community  0 points – cohort is based on selected group or not representative or no description of the cohort of interest |
| 2) Selection of the non-exposed cohort | 1 point – drawn from the same community as exposed cohort  0 points – drawn from a different source or no description provided |
| 3) Ascertainment of exposure | 1 point – exposure assessed prospectively or assigned from the medical record  0 points – retrospective assessment or may be at risk for recall bias or no description |
| 4) Demonstration that outcome of interest was not present at start of study | 1 point – exposure is assessed during pregnancy or before outcome can be present  0 points – exposure is assessed after outcome can be present |
| **Comparability** | |
| 5) Comparability of cohorts on the basis of the design or analysis | 1 point – study controls for potential confounder other than sociodemographic factors (such as social class, education, maternal age and ethnicity, child gender)  2 points – study controls for any other additional factor  0 points – no confounders included, or study does not control for sociodemographic factors |
| **Outcome** | |
| 6) Assessment of outcome | 1 point – record linkage or assessment based on multiple sources and/or evaluation by clinician  0 points - self-report or no reference to records or no assessment by clinician |
| 7) Was follow-up long enough for outcome to occur | 1 point – child age at least 6-7 years or 50% of the sample is older than 6 years  0 points – child age less than 6 years or >50% of the sample is younger than 6 years |
| 8) Adequacy of follow up of cohorts | 1 point – complete follow up or loss to follow-up is <50%  0 points - >50% of the sample lost to follow up or no information provided |

**Newcastle Ottawa Scale for case-control studies**

| **Item** | **Points** |
| --- | --- |
| **Selection** | |
| 1) Is the case definition adequate? | 1 point – independent validation (record linkage or using multiple sources and clinical assessment)  0 points – based on self-reports with no clinical validation or no description |
| 2) Representativeness of the cases | 1 point – eligible cases over a defined period of time, in a defined catchment area or clearly defined group  0 points – potential for selection bias or not stated |
| 3) Selection of controls | 1 point – same community as cases  0 points – hospital controls or not representing controls without mental health problems |
| 4) Definition of controls | 1 point – no history of disease (endpoint)  0 points – not meeting criteria for endpoint or no description |
| **Comparability** | |
| 5) Comparability of cases and controls on the basis of the design or analysis | 1 point – study controls for potential confounder other than sociodemographic factors (such as social class, education, maternal age and ethnicity, child gender)  2 points – study controls for any other additional factor  0 points – no confounders included, or study does not control for sociodemographic factors |
| **Exposure** | |
| 6) Ascertainment of exposure | 1 point – secure record, prospective measure or structured interview blind to case/control status  0 points – assessment not blinded to case/control status or retrospective self-report or may be at risk for recall bias or no description |
| 7) Same method of ascertainment for cases and controls | 1 point – Yes  0 points – No |
| 8) Non-response rate | 1 point – same rate for both groups  0 points – rate different or non-respondents described |

### Supplementary Table S1. Risk of bias assessment scores based on NOS scale of cohort, longitudinal and cross-sectional studies

| **Study** | **Item1** | **Item2** | **Item3** | **Item4** | **Item5** | **Item6** | **Item7** | **Item8** | **Total** |
| --- | --- | --- | --- | --- | --- | --- | --- | --- | --- |
| **Smoking** | | | | | | | | | |
| Ball et al., 2010 | 1 | 1 | 0 | 1 | 1 | 0 | 1 | 0 | **5** |
| Gustavson et al., 2017 | 1 | 1 | 1 | 1 | 2 | 1 | 1 | 1 | **9** |
| Langley et al., 2012 | 1 | 1 | 1 | 1 | 1 | 0 | 1 | 0 | **6** |
| Obel et al., 2011 | 1 | 1 | 1 | 1 | 1 | 1 | 1 | 1 | **8** |
| Obel et al., 2016 | 1 | 1 | 1 | 1 | 0 | 1 | 1 | 1 | **7** |
| Skoglund et al., 2014 | 1 | 1 | 1 | 1 | 0 | 1 | 1 | 1 | **7** |
| Zhu et al., 2014 | 1 | 1 | 1 | 1 | 2 | 1 | 1 | 1 | **9** |
| Lehn et al., 2007* | 1 | 1 | 1 | 1 | 1 | 0 | 1 | 1 | **7** |
| Lindblad et al., 2010 | 1 | 1 | 1 | 1 | 2 | 1 | 1 | 1 | **9** |
| Braun et al., 2006 | 1 | 1 | 0 | 0 | 1 | 0 | 1 | 0 | **4** |
| Wakschlag et al., 1997 | 0 | 1 | 0 | 0 | 2 | 1 | 1 | 1 | **6** |
| Talati et al., 2016* | 0 | 1 | 0 | 0 | 0 | 1 | 1 | 1 | **4** |
| Braun et al., 2008 | 1 | 1 | 0 | 0 | 0 | 0 | 1 | 1 | **4** |
| Ellis et al., 2012 | 1 | 1 | 0 | 0 | 2 | 0 | 0 | 1 | **5** |
| Nigg et al., 2007 | 1 | 1 | 0 | 0 | 1 | 0 | 1 | 1 | **5** |
| Talati et al., 2017 | 0 | 1 | 0 | 1 | 0 | 1 | 1 | 1 | **5** |
| Weissman et al., 1999 | 0 | 1 | 0 | 0 | 0 | 1 | 1 | 1 | **4** |
| Nomura et al., 2010 | 0 | 0 | 0 | 0 | 2 | 1 | 0 | 0 | **3** |
| Neuman et al., 2007 | 1 | 1 | 0 | 1 | 2 | 0 | 1 | 0 | **6** |
| Koshy et al., 2011 | 0 | 1 | 0 | 0 | 0 | 0 | 1 | 0 | **2** |
| Sciberras et al., 2011 | 1 | 1 | 0 | 0 | 1 | 0 | 1 | 1 | **5** |
| Froehlich et al., 2009 | 1 | 1 | 0 | 0 | 1 | 0 | 1 | 0 | **4** |
| Knopik et al., 2005 | 1 | 1 | 0 | 0 | 2 | 0 | 1 | 1 | **6** |
| Knopik et al., 2006 | 1 | 1 | 0 | 0 | 1 | 0 | 1 | 0 | **4** |
| Schwenke et al., 2018 | 0 | 1 | 1 | 0 | 0 | 0 | 1 | 0 | **3** |
| **Alcohol** | | | | | | | | | |
| Eilertsen et al., 2017 | 1 | 1 | 1 | 1 | 1 | 1 | 0 | 0 | **6** |
| Larkby et al., 2011 | 1 | 1 | 1 | 1 | 2 | 0 | 1 | 1 | **8** |
| Lees et al., 2020 | 1 | 1 | 0 | 0 | 2 | 0 | 1 | 1 | **6** |
| Mitchell et al., 2020 | 1 | 1 | 0 | 1 | 1 | 0 | 1 | 1 | **6** |
| Pagnin et al., 2019 | 0 | 1 | 1 | 1 | 0 | 0 | 1 | 0 | **4** |
| Weile et al., 2020 | 1 | 1 | 1 | 1 | 2 | 1 | 1 | 0 | **8** |
| **Caffeine** | | | | | | | | | |
| Del-Ponte et al., 2016 | 1 | 1 | 0 | 1 | 2 | 1 | 1 | 1 | **8** |
| Linnet et al., 2008 | 1 | 1 | 1 | 1 | 2 | 1 | 1 | 1 | **9** |
| **Smoking and Alcohol** | | | | | | | | | |
| Fergusson et al., 1998 | 1 | 1 | 1 | 1 | 2 | 0 | 1 | 1 | **8** |
| Pohlabeln et al., 2017 | 0 | 1 | 0 | 0 | 2 | 0 | 1 | 1 | **5** |
| Sagiv et al., 2013 | 1 | 1 | 1 | 1 | 2 | 1 | 1 | 1 | **9** |
| Schmitt et al., 2012 | 1 | 1 | 0 | 0 | 2 | 0 | 1 | 0 | **5** |
| Whitbeck et al., 2009 | 0 | 1 | 0 | 0 | 0 | 0 | 1 | 0 | **2** |
| **Smoking, alcohol and caffeine** | | | | | | | | | |
| Russell et al., 2015 | 0 | 1 | 0 | 0 | 0 | 0 | 1 | 0 | **2** |

### Supplementary Table S2. Risk of bias assessment scores based on NOS scale of case-control studies

| **Study** | **Item1** | **Item2** | **Item3** | **Item4** | **Item5** | **Item6** | **Item7** | **Item8** | **Total** |
| --- | --- | --- | --- | --- | --- | --- | --- | --- | --- |
| **Smoking** | | | | | | | | | |
| Biederman et al., 2009 | 1 | 1 | 0 | 1 | 2 | 0 | 1 | 0 | **6** |
| Gard et al., 2010 | 0 | 0 | 0 | 0 | 1 | 0 | 1 | 0 | **2** |
| Gustaffson and Kallen, 2010 | 1 | 1 | 1 | 1 | 0 | 1 | 1 | 1 | **7** |
| Joelsson et al., 2016 | 1 | 1 | 1 | 1 | 2 | 1 | 1 | 1 | **9** |
| Linnet et al., 2005 | 1 | 1 | 1 | 1 | 2 | 1 | 1 | 1 | **9** |
| Milberger et al., 1996 | 1 | 1 | 0 | 1 | 1 | 0 | 1 | 0 | **5** |
| Milberger et al., 1998 | 1 | 1 | 0 | 1 | 1 | 0 | 1 | 0 | **5** |
| Motlagh et al., 2010 | 1 | 1 | 1 | 1 | 0 | 0 | 0 | 0 | **4** |
| Schmitz et al., 2006 | 1 | 1 | 1 | 1 | 2 | 0 | 1 | 1 | **8** |
| Silva et al., 2014 | 1 | 1 | 1 | 1 | 2 | 0 | 1 | 1 | **8** |
| Todd et al., 2007 | 0 | 1 | 1 | 1 | 1 | 0 | 1 | 0 | **5** |
| Yoshimasu et al., 2009 | 1 | 1 | 0 | 1 | 2 | 0 | 1 | 1 | **7** |
| Altink et al., 2009 | 1 | 1 | 1 | 1 | 2 | 0 | 1 | 0 | **7** |
| Altink et al., 2008 | 1 | 1 | 1 | 1 | 0 | 0 | 1 | 0 | **5** |
| Arnold et al., 2005 | 1 | 1 | 1 | 1 | 2 | 0 | 1 | 0 | **7** |
| Biederman et al., 2017 | 1 | 1 | 0 | 1 | 0 | 0 | 1 | 0 | **4** |
| Oerlemans et al., 2016 | 1 | 1 | 1 | 1 | 0 | 0 | 1 | 0 | **5** |
| Wang et al., 2019 | 1 | 1 | 1 | 1 | 1 | 0 | 1 | 0 | **6** |
| Sourander et al., 2019 | 1 | 1 | 1 | 1 | 1 | 1 | 1 | 1 | **8** |
| **Smoking and Alcohol** | | | | | | | | | |
| Ketzer et al., 2012 | 1 | 1 | 1 | 1 | 1 | 0 | 1 | 0 | **6** |
| Mick et al., 2002 | 1 | 1 | 0 | 1 | 1 | 0 | 1 | 0 | **5** |
| Pineda et al., 2007 | 1 | 1 | 1 | 1 | 0 | 0 | 1 | 1 | **6** |
| Wiggs et al., 2016 | 1 | 1 | 1 | 1 | 1 | 0 | 1 | 0 | **6** |
| **Alcohol and caffeine** | | | | | | | | | |
| Kim et al., 2009 | 0 | 1 | 1 | 1 | 0 | 0 | 1 | 0 | **4** |

### Supplementary Table S3. List of studies excluded

|  | **Author** | **Year** | **Title** | **Exclusion reason** |
| --- | --- | --- | --- | --- |
| 1. | Brookes et al. | 2006 | A common haplotype of the dopamine transporter gene associated with attention-deficit/hyperactivity disorder and interacting with maternal use of alcohol during pregnancy | no comparison/control group |
| 2. | Freitag et al. | 2012 | Biological and psychosocial environmental risk factors influence symptom severity and psychiatric comorbidity in children with ADHD | no comparison/control group |
| 3. | Brookes et al. | 2006 | Association of Fatty Acid Desaturase Genes with Attention-Deficit/Hyperactivity Disorder | no comparison/control group |
| 4. | Langley et al. | 2007 | Effects of low birth weight, maternal smoking in pregnancy and social class on the phenotypic manifestation of Attention Deficit Hyperactivity Disorder and associated antisocial behaviour: investigation in a clinical sample | no comparison/control group |
| 5. | Langley et al. | 2008 | Testing for gene x environment interaction effects in attention deficit hyperactivity disorder and associated antisocial behavior | no comparison/control group |
| 6. | Thakur et al. | 2012 | Comprehensive Phenotype/Genotype Analyses of the Norepinephrine Transporter Gene (SLC6A2) in ADHD: Relation to Maternal Smoking during Pregnancy | no comparison/control group |
| 7. | Thakur et al. | 2013 | Maternal smoking during pregnancy and ADHD: A comprehensive clinical and neurocognitive characterization | no comparison/control group |
| 8. | Biederman et al. | 2012 | Does exposure to maternal smoking during pregnancy affect the clinical features of ADHD? Results from a controlled study | no comparison/control group |
| 9. | Bhatara et al. | 2006 | Association of attention deficit hyperactivity disorder and gestational alcohol exposure: An exploratory study | no comparison/control group |
| 10. | Xu et al. | 2010 | Racial differences in the effects of postnatal environmental tobacco smoke on neurodevelopment | wrong exposure |
| 11. | Mulligan et al. | 2013 | Home environment: association with hyperactivity/impulsivity in children with ADHD and their non-ADHD siblings | wrong exposure |
| 12. | Naeye and Peters | 1984 | Mental development of children whose mothers smoked during pregnancy | wrong outcome |
| 13. | Holz et al. | 2014 | Effect of Prenatal Exposure to Tobacco Smoke on Inhibitory Control Neuroimaging Results from a 25-Year Prospective Study | wrong outcome |
| 14. | Martel and Roberts | 2014 | Prenatal testosterone increases sensitivity to prenatal stressors in males with disruptive behavior disorders | wrong exposure |
| 15. | Wakschlag and Keenan | 2001 | Clinical significance and correlates of disruptive behavior in environmentally at-risk preschoolers | don' t meet outcome criteria  (maternal reported conduct disorder and ADHD symptoms) |
| 16. | Sengupta et al. | 2015 | Parental psychopathology in families of children with attention-deficit/hyperactivity disorder and exposed to maternal smoking during pregnancy | wrong outcome |
| 17. | Mick et al. | 2002 | Impact of low birth weight on attention-deficit hyperactivity disorder | wrong exposure |
| 18. | Grabell and Olson | 2010 | Executive functioning as a mediating factor of prenatal alcohol exposure and externalizing problems in preschool children | conference/meeting abstract |
| 19. | Schmitz et al. | 2017 | Pre- and perinatal risk factors in autism spectrum disorder and attention deficit/hyperactivity disorder | not English language |
| 20. | Willoughby et al. | 2012 | Parent-Reported Attention Deficit/Hyperactivity Symptomatology in Preschool-Aged Children: Factor Structure, Developmental Change, and Early Risk Factors | no comparison/control group |
| 21. | Wiliams et al. | 1998 | Maternal cigarette smoking and child psychiatric morbidity: a longitudinal study | wrong outcome |
| 22. | Whitaker et al. | 2011 | Serial pediatric symptom checklist screening in children with prenatal drug exposure | wrong exposure |
| 23. | Whitaker et al. | 2006 | Food insecurity and the risks of depression and anxiety in mothers and behavior problems in their preschool-aged children | wrong exposure |
| 24. | Way and Rojahn | 2012 | Psycho-social characteristics of children with prenatal alcohol exposure, compared to children with Down syndrome and typical children | wrong outcome |
| 25. | Twardella et al. | 2010 | Exposure to secondhand tobacco smoke and child behaviour - results from a cross-sectional study among preschool children in Bavaria | wrong exposure |
| 26. | Vuijk et al. | 2006 | Prenatal smoking predicts non-responsiveness to an intervention targeting attention-deficit/hyperactivity symptoms in elementary schoolchildren | retracted |
| 27. | Van Den Berg and Marcoen | 2004 | High antenatal maternal anxiety is related to ADHD symptoms, externalizing problems, and anxiety in 8- and 9-year-olds | wrong exposure |
| 28. | Todd and Neuman | 2007 | Gene-environment interactions in the development of combined type ADHD: evidence for a synapse-based model | duplicate |
| 29. | Tiesler and Heinrich | 2014 | Prenatal nicotine exposure and child behavioural problems | review |
| 30. | Teramoto et al. | 2005 | Problematic behaviours of 3-year-old children in Japan: Relationship with socioeconomic and family backgrounds | externalising disorder not specified |
| 31. | Tearne et al. | 2015 | The association between prenatal environment and children's mental health trajectories from 2 to 14 years | externalising disorder not specified |
| 32. | Höök et al. | 2006 | Prenatal and postnatal maternal smoking as risk factors for preschool children's mental health | externalising disorder not specified |
| 33. | Schonfeld et al. | 2005 | Moral maturity and delinquency after prenatal alcohol exposure | FAS sample |
| 34. | Roza et al. | 2009 | Maternal smoking during pregnancy and child behaviour problems: the Generation R Study | externalising disorder not specified |
| 35. | Schlotz et al. | 2010 | Lower maternal folate status in early pregnancy is associated with childhood hyperactivity and peer problems in offspring | wrong exposure |
| 36. | Salom et al. | 2015 | Familial factors associated with development of alcohol and mental health comorbidity | wrong exposure  (not measured during pregnancy) |
| 37. | Salom et al. | 2016 | Predictors of comorbid polysubstance use and mental health disorders in young adults-a latent class analysis | wrong outcome |
| 38. | Ruchkin et al. | 2008 | Developmental pathway modeling in considering behavior problems in young Russian children | externalising disorder not specified |
| 39. | Pauli-Rott et al. | 2017 | Psychosocial risk factors underlie the link between attention deficit hyperactivity symptoms and overweight at school entry | wrong exposure |
| 40. | Paley et al. | 2005 | Prenatal Alcohol Exposure, Child Externalizing Behavior, and Maternal Stress | externalising disorder not specified |
| 41. | Oulhote and Bouchard | 2013 | Urinary metabolites of organophosphate and pyrethroid pesticides and behavioral problems in Canadian children | wrong exposure |
| 42. | Olson et al. | 1992 | Prenatal exposure to alcohol and school problems in late childhood: A longitudinal prospective study | externalising disorder not specified |
| 43. | Robinson et al. | 2010 | Low-moderate prenatal alcohol exposure and risk to child behavioural development: A prospective cohort study | externalising disorder not specified |
| 44. | Robinson et al. | 2008 | Pre- and postnatal influences on preschool mental health: A large-scale cohort study | externalising disorder not specified |
| 45. | Robinson et al. | 2010 | Smoking cessation in pregnancy and the risk of child behavioural problems: A longitudinal prospective cohort study | externalising disorder not specified |
| 46. | Pineda et al. | 2003 | Perinatal factors associated with attention deficit/hyperactivity diagnosis in Colombian Paisa children] | not English language |
| 47. | Pfinder et al. | 2014 | Impact of Moderate Prenatal Alcohol Exposure on Problem Behaviors in Preschool and School Children | duplicate |
| 48. | Petkovsek et al. | 2014 | Prenatal smoking and genetic risk: Examining the childhood origins of externalizing behavioral problems | externalising disorder not specified |
| 49. | Oćallaghan et al. | 1997 | Obstetric and perinatal factors as predictors of child behaviour at 5 years | externalising disorder not specified |
| 50. | O'Brien et al. | 2013 | Do dopamine gene variants and prenatal smoking interactively predict youth externalizing behavior? | no comparison/control group |
| 51. | Niemela et al. | 2016 | Maternal smoking during pregnancy and offpsring's psychiatric morbidity in early adulthood. Findings from the Finnish Family Competence Birth Cohort Study | conference/meeting abstract |
| 52. | Niclasen et al. | 2012 | Prenatal exposure to alcohol and neurobehavioural development at age seven | conference/meeting abstract |
| 53. | Najman et al. | 2000 | Preschool children and behaviour problems - A prospective study | externalising disorder not specified |
| 54. | Glass et al. | 2014 | Correspondence of parent report and laboratory measures of inattention and hyperactivity in children with heavy prenatal alcohol exposure | FAS sample |
| 55. | Fryer et al. | 2007 | Evaluation of psychopathological conditions in children with heavy prenatal alcohol exposure | FAS sample |
| 56. | Mina et al. | 2017 | Prenatal exposure to very severe maternal obesity is associated with adverse neuropsychiatric outcomes in children | wrong exposure |
| 57. | Melchior et al. | 2012 | Food Insecurity and Children's Mental Health: A Prospective Birth Cohort Study | wrong exposure |
| 58. | Mullen et al. | 2017 | Effects of prenatal alcohol exposure on child behaviour outcomes at age five: Lifeways Cross-Generational Cohort Study | conference/meeting abstract |
| 59. | Monshouwer et al | 2011 | Prenatal smoking exposure and the risk of behavioral problems and substance use in adolescence: The TRAILS study | externalising disorder not specified |
| 60. | Minnes et al. | 2010 | The effects of prenatal cocaine exposure on problem behavior in children 4-10 years | wrong exposure |
| 61. | Momany et al. | 2017 | Sex moderates the impact of birth weight on child externalizing psychopathology | wrong exposure |
| 62. | Milberger et al. | 1996 | Is maternal smoking during pregnancy a risk factor for attention deficit hyperactivity disorder in children? | duplicate |
| 63. | MacKinnon et al. | 2018 | The Association Between Prenatal Stress and Externalizing Symptoms in Childhood: Evidence From the Avon Longitudinal Study of Parents and Children | wrong exposure |
| 64. | Lukkari et al. | 2012 | Exposure to Obstetric Complications in Relation to Subsequent Psychiatric Disorders of Adolescent Inpatients: Specific Focus on Gender Differences | wrong exposure |
| 65. | McLaughlin et al. | 2011 | Caregiver and self-report of mental health symptoms in 9-year old children with prenatal cocaine exposure | wrong exposure |
| 66. | McCrory et al. | 2012 | Prenatal exposure to maternal smoking and childhood behavioural problems: A quasi-experimental approach | externalising disorder not specified |
| 67. | Maughan et al. | 2004 | Prenatal smoking and early childhood conduct problems: Testing genetic and environmental explanations of the association | don't meet outcome criteria  (maternal and teacher reported measure of conduct disorder and delinquent behaviour symptoms) |
| 68. | Marceau et al. | 2018 | Within-Family Effects of Smoking during Pregnancy on ADHD: the Importance of Phenotype | don't meet outcome criteria  (maternal reported ADHD symptoms) |
| 69. | Linnet et al. | 2003 | Maternal Lifestyle Factors in Pregnancy Risk of Attention Deficit Hyperactivity Disorder and Associated Behaviors: Review of the Current Evidence | review |
| 70. | Langley et al. | 2005 | Maternal smoking during pregnancy as an environmental risk factor for attention deficit hyperactivity disorder behaviour. A review | review |
| 71. | Coles et al. | 1997 | A comparison of children affected by prenatal alcohol exposure and attention deficit, hyperactivity disorder | FAS sample |
| 72. | Walthall et al. | 2008 | A comparison of psychopathology in children with and without prenatal alcohol exposure | FAS sample |
| 73. | Neiderhiser et al. | 2016 | Estimating the Roles of Genetic Risk, Perinatal Risk, and Marital Hostility on Early Childhood Adjustment: Medical Records and Self-Reports | externalising disorder not specified |
| 74. | Max et al. | 2013 | Attention deficit hyperactivity disorder among children exposed to secondhand smoke: A logistic regression analysis of secondary data | wrong exposure |
| 75. | Marin-Mendez et al. | 2015 | Environmental factors associated with suspected ADHD in preschoolers using a screening tool (ADHD-RS-IV-P) | wrong exposure  (not measured in pregnancy) |
| 76. | Maat et al. | 2017 | Attention-deficit hyperactivity disorder (adhd) symptoms in children adopted from poland and their atypical association patterns: A bayesian approach | no exposure-outcome association measured |
| 77. | Linnet al. | 2006 | Gestational age, birth weight, and the risk of hyperkinetic disorder | wrong exposure |
| 78. | LaGasse et al. | 2012 | Prenatal methamphetamine exposure and childhood behavior problems at 3 and 5 years of age | wrong exposure |
| 79. | Lewis et al. | 2016 | Prospective Memory Impairment in Children with Prenatal Alcohol Exposure | FAS sample |
| 80. | Lahti et al. | 2006 | Small body size at birth and behavioural symptoms of ADHD in children aged five to six years | wrong exposure |
| 81. | Maatta et al. | 2017 | Maternal Smoking During Pregnancy Is Associated With Offspring's Musculoskeletal Pain in Adolescence: Structural Equation Modeling | wrong outcome |
| 82. | Li et al. | 1996 | Study of child's behavior problem in logistic regression analysis | not English language |
| 83. | Langley et al. | 2010 | Maternal and paternal smoking during pregnancy and risk for child ADHD: A test for causal associations | conference/meeting abstract |
| 84. | Kukla et al. | 2006 | Smoking of mothers during pregnancy in relation to mental and motoric development disorders in 4- and 5-year-old children. The elspac study results | not English language |
| 85. | Wakschlag et al. | 2006 | Elucidating early mechanisms of developmental psychopathology: The case of prenatal smoking and disruptive behavior | wrong outcome |
| 86. | Leung et al. | 2015 | Early second-hand smoke exposure and child and adolescent mental health: Evidence from Hong Kong's 'Children of 1997' birth cohort | wrong exposure |
| 87. | Silva et al. | 2015 | Comorbidities of Attention Deficit Hyperactivity Disorder: Pregnancy Risk Factors and Parent Mental Health | combined outcome |
| 88. | Nanson and Hiscock | 1990 | Attention deficits in children exposed to alcohol prenatally | FAS sample |
| 89. | Marino et al. | 2010 | Association of maternal lifestyle factors in pregnancy and birth weight with attention-deficit/hyperactivity disorder in a population of Italian children | conference/meeting abstract |
| 90. | Rasmussen et al. | 2011 | An evaluation of social skills in children with and without prenatal alcohol exposure | FAS sample |
| 91. | Tervo et al. | 2017 | A Prospective 30-Year Follow-Up of ADHD Associated With Perinatal Risks | wrong exposure |
| 92. | Wiggs et al. | 2017 | A Family-Based Study of the Association Between Labor Induction and Offspring Attention-Deficit Hyperactivity Disorder and Low Academic Achievement | wrong exposure |
| 93. | Knudsen et al. | 2014 | Maternal pre-pregnancy risk drinking and toddler behavior problems: the Norwegian Mother and Child Cohort Study | externalising disorder not specified |
| 94. | Knopik et al. | 2010 | Smoking during pregnancy, maternal xenobiotic metabolism genes, and child externalizing behavior: A case-crossover design | conference/meeting abstract |
| 95. | Knopik et al. | 2009 | Maternal smoking during pregnancy and child outcomes: Real or spurious effect? | review |
| 96. | Knopik et al. | 2015 | Smoking during pregnancy and ADHD risk: A genetically-informed, multiple-rater approach | duplicate |
| 97. | Keyes et al. | 2012 | Associations of prenatal maternal smoking with offspring hyperactivity: Causal or confounded? | duplicate |
| 98. | Kesmodel et al. | 2010 | Lifestyle during pregnancy: neurodevelopmental effects at 5 years of age. The design and implementation of a prospective follow-up study | wrong outcome |
| 99. | Katwan et al. | 2011 | Childhood behavioural and developmental disorders: association with maternal alcohol consumption in Cape Town, South Africa | externalising disorder not specified |
| 100. | Kahn et al. | 2005 | Intergenerational health disparities: Socioeconomic status, women's health conditions, and child behavior problems | externalising disorder not specified |
| 101. | Johnson et al. | 2001 | Moderate alcohol and tobacco use during pregnancy and child behavior outcomes | don't meet outcome criteria  (maternal reported measure of conduct disorder) |
| 102. | Jaspers et al. | 2012 | Trajectories of psychosocial problems in adolescents predicted by findings from early well-child assessments | externalising disorder not specified |
| 103. | Jaspers et al. | 2010 | Early findings of preventive child healthcare professionals predict psychosocial problems in preadolescence: The TRAILS study | externalising disorder not specified |
| 104. | Jacka et al. | 2013 | Maternal and early postnatal nutrition and mental health of offspring by age 5 years: A prospective cohort study | wrong exposure |
| 105. | Irner et al. | 2014 | Cognitive, emotional and social development in adolescents born to substance using women | wrong exposure |
| 106. | Holz et al. | 2015 | The long-term impact of early life poverty on orbitofrontal cortex volume in adulthood: Results from a prospective study over 25 years | wrong exposure |
| 107. | Infante et al. | 2015 | Objective assessment of ADHD core symptoms in children with heavy prenatal alcohol exposure | FAS sample |
| 108. | Huizink et al. | 2006 | Maternal smoking, drinking or cannabis use during pregnancy and neurobehavioral and cognitive functioning in human offspring | review |
| 109. | Huang et al. | 2018 | Maternal smoking and attention-deficit/hyperactivity disorder in offspring: A meta-analysis | review/meta-analysis |
| 110. | Henry et al. | 2007 | Neurobiology and neurodevelopmental impact of childhood traumatic stress and prenatal alcohol exposure | FAS sample |
| 111. | Habek et al. | 2011 | Adverse pregnancy outcomes and long-term morbidity after early fetal hypokinesia in maternal smoking pregnancies | no exposure-outcome association measured |
| 112. | Gray et al. | 2004 | Prevalence, stability, and predictors of clinically significant behavior problems in low birth weight children at 3, 5, and 8 years of age | externalising disorder not specified |
| 113. | Hay et al. | 2008 | Antepartum and postpartum exposure to maternal depression: Different effects on different adolescent outcomes | wrong exposure |
| 114. | deMaat et al. | 2018 | Attention-Deficit Hyperactivity Disorder (ADHD) Symptoms in Children Adopted from Poland and their Atypical Association Patterns: a Bayesian Approach | duplicate |
| 115. | Burden et al. | 2010 | An event-related potential study of response inhibition in ADHD with and without prenatal alcohol exposure | wrong outcome |
| 116. | Gronimus et al. | 2009 | Maternal alcohol consumption | review |
| 117. | Griesler et al. | 1998 | Maternal smoking in pregnancy, child behavior problems, and adolescent smoking | externalising disorder not specified |
| 118. | Graham et al. | 2013 | Prenatal Alcohol Exposure, Attention-Deficit/Hyperactivity Disorder, and Sluggish Cognitive Tempo | FAS sample |
| 119. | Goodwin et al. | 2013 | Maternal nicotine dependence and prenatal smoking and their association with mental disorders among offspring from the community | conference/meeting abstract |
| 120. | Goldman et al. | 2011 | Direct and modifying influences of selected risk factors on children's pre-adoption functioning and post-adoption adjustment | wrong outcome |
| 121. | Glass et al. | 2013 | Neuropsychological deficits associated with heavy prenatal alcohol exposure are not exacerbated by ADHD | FAS sample |
| 122. | Amiri et al. | 2012 | Pregnancy-related maternal risk factors of attention-deficit hyperactivity disorder: a case-control study | combined exposure (tobacco and alcohol) |
| 123. | Fitzpatrick et al. | 2017 | Prevalence and profile of Neurodevelopment and Fetal Alcohol Spectrum Disorder (FASD) amongst Australian Aboriginal children living in remote communities | FAS sample |
| 124. | Furtado and Roriz | 2016 | Inattention and impulsivity associated with prenatal alcohol exposure in a prospective cohort study with 11-years-old Brazilian children | wrong outcome |
| 125. | Furlong et al. | 2017 | Early life characteristics and neurodevelopmental phenotypes in the mount sinai children's environmental health center | externalising disorder not specified |
| 126. | Froehlich et al. | 2011 | Update on environmental risk factors for attention-deficit/hyperactivity disorder | review |
| 127. | Flak et al. | 2014 | The association of mild, moderate, and binge prenatal alcohol exposure and child neuropsychological outcomes: A meta-analysis | review/meta-analysis |
| 128. | Finnegan | 1981 | The effects of narcotics and alcohol on pregnancy and the newborn | review |
| 129. | Erb and Andresen | 1981 | Hyperactivity: a possible consequence of maternal alcohol consumption | review |
| 130. | Ekblad et al. | 2017 | Maternal smoking during pregnancy and the risk of psychiatric morbidity in singleton sibling pairs | externalising disorder not specified |
| 131. | Aronson et al. | 1997 | Attention deficits and autistic spectrum problems in children exposed to alcohol during gestation: A follow-up study | FAS sample |
| 132. | Eiden et al. | 2011 | Child behavior problems among cocaine-exposed toddlers: Indirect and interactive effects | wrong exposure |
| 133. | Camprodon-Rosanas et al. | 2017 | Sluggish Cognitive Tempo: Sociodemographic, Behavioral, and Clinical Characteristics in a Population of Catalan School Children | wrong outcome |
| 134. | Delaney-Black et al. | 1998 | Prenatal Coke: What's Behind the Smoke?: Prenatal Cocaine/Alcohol Exposure and School-Age Outcomes: The SCHOO-BE Experiencea | no exposure-outcome association measured |
| 135. | Button et al. | 2007 | The relationship of maternal smoking to psychological problems in the offspring | review |
| 136. | Boseck et al. | 2015 | Cognitive and Adaptive Skill Profile Differences in Children With Attention-Deficit Hyperactivity Disorder With and Without Comorbid Fetal Alcohol Spectrum Disorder | FAS sample |
| 137. | Biederman et al. | 2014 | Is ADHD a risk for posttraumatic stress disorder (PTSD)? Results from a large longitudinal study of referred children with and without ADHD | wrong outcome |
| 138. | Boutwell et al. | 2011 | Prenatal exposure to cigarette smoke and childhood externalizing behavioral problems: A propensity score matching approach | externalising disorder not specified |
| 139. | Boutwell and Beaver | 2010 | Maternal cigarette smoking during pregnancy and offspring externalizing behavioral problems: A propensity score matching analysis | externalising disorder not specified |
| 140. | Bor et al. | 1997 | The relationship between low family income and psychological disturbance in young children: An Australian longitudinal study | externalising disorder not specified |
| 141. | Beal et al. | 2015 | Associations between the prenatal environment and cardiovascular risk factors in adolescent girls: Internalizing and externalizing behavior symptoms as mediators | wrong outcome |
| 142. | Basgul et al. | 2011 | Frequency and correlates of psychiatric disorders in early childhood: a study of population and clinical samples in Turkey | no exposure-outcome association measured |
| 143. | Barr et al. | 2006 | Binge Drinking During Pregnancy as a Predictor of Psychiatric Disorders on the Structured Clinical Interview for DSM-IV in Young Adult Offspring | wrong outcome |
| 144. | Bailey et al. | 2005 | Gender and alcohol moderate prenatal cocaine effects on teacher-report of child behavior | wrong exposure |
| 145. | Bada et al. | 2007 | Impact of prenatal cocaine exposure on child behavior problems through school age | externalising disorder not specified |
| 146. | Autti-Ramo | 2000 | Twelve-year follow-up of children exposed to alcohol in utero | FAS sample |
| 147. | Ashford et al. | 2008 | Prenatal smoking and internalizing and externalizing problems in children studied from childhood to late adolescence | don't meet outcome criteria  (maternal reported ADHD symptoms) |
| 148. | Arbuckle et al. | 2016 | Bisphenol A, phthalates and lead and learning and behavioral problems in Canadian children 6-11 years of age: CHMS 2007-2009 | wrong exposure |
| 149. | Yao | 2017 | Mother Smoking During Pregnancy and ADHD in Children | conference/meeting abstract |
| 150. | Gaysina et al. | 2013 | Prenatal active or passive tobacco smoke exposure and the risk of child externalising problems: Evidence from a genetically-sensitive research design | conference/meeting abstract |
| 151. | Gaysina et al. | 2012 | Maternal smoking during pregnancy and offspring conduct problems: The evidence for the association using genetically-sensitive designs | conference/meeting abstract |
| 152. | Fago | 2013 | Impact of prenatal alcohol exposure and pre-adoption placement on school-age functioning of intercountry-adopted children | dissertation |
| 153. | Bidwell et al. | 2015 | A propensity scoring approach to characterizing the effects of maternal smoking during pregnancy on initial responses to tobacco and alcohol in adolescents | conference/meeting abstract |
| 154. | Baler et al. | 2008 | Is fetal brain monoamine oxidase inhibition the missing link between maternal smoking and conduct disorders? | review |
| 155. | Wolke et al. | 2011 | The impact of light drinking in pregnancy on children's behavioural and cognitive development | conference/meeting abstract |
| 156. | Fried et al. | 1995 | Prenatal exposure to marihuana and tobacco during infancy, early and middle childhood: effects and an attempt at synthesis | conference/meeting abstract |
| 157. | Freitag et al. | 2015 | Pre-and perinatal risk factors in attention-deficit/ hyperactivity disorder and autism spectrum disorders | conference/meeting abstract |
| 158. | Fitzpatrick and Pagani | 2011 | Compelling evidence that child impulsivity in fourth grade is predicted by maternal smoking during pregnancy | conference/meeting abstract |
| 159. | Evrensel et al. | 2015 | Rate of perinatal nicotine exposure in children with the diagnosis of Attention Deficit Hyperactivity Disorder | conference/meeting abstract |
| 160. | Estabrook et al. | 2014 | The importance of reliable longitudinal measurement in the assessment of prenatal smoking and its relation to youth externalizing behavior | conference/meeting abstract |
| 161. | Enoch et al. | 2014 | A longitudinal study in mothers and firstborn children of genetic and environmental influences on externalizing and internalizing disorders across development | conference/meeting abstract |
| 162. | Enoch et al. | 2015 | A prospective cohort study of influences on internalizing and externalizing behaviors across childhood | conference/meeting abstract |
| 163. | Dodge et al. | 2010 | Protective effects of the alcohol dehydrogenase-adh1b*3 allele in adolescents exposed to alcohol during pregnancy | conference/meeting abstract |
| 164. | Coles et al. | 2014 | Prenatal tobacco exposure and behavior regulation at 24 months: Child language and maternal psychological symptoms as moderators | conference/meeting abstract |
| 165. | Brookes et al. | 2005 | A common haplotype of the dopamine transporter gene is associated with attention deficit hyperactivity disorder and interacts with prenatal exposure to alcohol | conference/meeting abstract |
| 166. | Bilgec et al. | 2013 | Possible prenatal and genetic factors in the etiology of attention deficit hyperactivity disorder: a Turkish referred sample | conference/meeting abstract |
| 167. | Bazinet etal. | 2010 | Motor activity and inattention during a sustained attention task in children with heavy prenatal alcohol exposure | conference/meeting abstract |
| 168. | Adnams et al. | 2014 | Behavior in secondary school learnerswith prenatal alcohol exposure in south Africa | conference/meeting abstract |
| 169. | McIntosh et al. | 1995 | Utilization of maternal perinatal risk indicators in the differential diagnosis of ADHD and UADD children | no exposure-outcome association measured |
| 170. | Jacobson et al. | 2011 | Number Processing in Adolescents With Prenatal Alcohol Exposure and ADHD: Differences in the Neurobehavioral Phenotype | wrong outcome |
| 171. | Hayatbakhsh et al. | 2011 | A longitudinal study of child mental health and problem behaviours at 14years of age following unplanned pregnancy | wrong exposure |
| 172. | Gaysina et al. | 2013 | Maternal smoking during pregnancy and offspring conduct problems: Evidence from 3 independent genetically sensitive research designs | don't meet outcome criteria  (maternal reported conduct disorder symptoms) |
| 173. | Yu et al. | 2016 | Attention Deficit/Hyperactivity Disorder and Urinary Nonylphenol Levels: A Case-Control Study in Taiwanese Children | wrong exposure |
| 174. | Gutvirtz et al. | 2018 | Maternal smoking and long-term neurological morbidity of the offspring | wrong outcome |
| 175. | Wakschlag and Hans | 2002 | Maternal smoking during pregnancy and conduct problems in high-risk youth: A developmental framework | don't meet outcome criteria  (maternal reported conduct disorder symptoms) |
| 176. | Ware et al. | 2013 | The effects of prenatal alcohol exposure and attention-deficit/hyperactivity disorder on psychopathology and behavior | FAS sample |
| 177. | Galera et al. | 2011 | Early risk factors for hyperactivity-impulsivity and inattention trajectories from age 17 months to 8 years | don't meet outcome criteria  (maternal reported ADHD symptoms) |
| 178. | Wasserman et al. | 2001 | Contribution of maternal smoking during pregnancy and lead exposure to early child behavior problems | don't meet outcome criteria  (maternal reported conduct disorder and ADHD symptoms) |
| 179. | Thapar et al. | 2009 | Prenatal smoking might not cause attention-deficit/hyperactivity disorder: Evidence from a novel design | don't meet outcome criteria  (maternal reported ADHD symptoms) |
| 180. | Wilson et al. | 2013 | What predicts persistent early conduct problems? Evidence from the Growing Up in Scotland cohort | don't meet outcome criteria  (maternal reported conduct disorder symptoms) |
| 181. | Weitzman et al. | 1992 | Maternal smoking and behavior problems of children | don't meet outcome criteria  (maternal reported ADHD symptoms) |
| 182. | Wakschlag et al. | 2006 | Is prenatal smoking associated with a developmental pattern of conduct problems in young boys? | don't meet outcome criteria  (maternal reported conduct disorder and ADHD symptoms) |
| 183. | Tiesler et al. | 2011 | Passive smoking and behavioural problems in children: results from the LISAplus prospective birth cohort study | don't meet outcome criteria  (maternal reported conduct disorder and ADHD symptoms) |
| 184. | Olson et al. | 1997 | Association of prenatal alcohol exposure with behavioral and learning problems in early adolescence | don't meet outcome criteria  (maternal reported conduct disorder symptoms) |
| 185. | Alvik et al. | 2013 | Early fetal binge alcohol exposure predicts high behavioral symptom scores in 5.5-year-old children | don't meet outcome criteria  (maternal reported conduct disorder, ADHD symptoms) |
| 186. | Thapar et al. | 2003 | Maternal smoking during pregnancy and attention deficit hyperactivity disorder symptoms in offspring | don't meet outcome criteria  (maternal reported ADHD symptoms) |
| 187. | Tanaka et al. | 2016 | Perinatal smoking exposure and behavioral problems in Japanese children aged 5 years: The Kyushu Okinawa Maternal and Child Health Study | don't meet outcome criteria  (maternal reported conduct disorder and ADHD symptoms) |
| 188. | Staroselsky et al. | 2009 | Both parental psychopathology and prenatal maternal alcohol dependency can predict the behavioral phenotype in children | don't meet outcome criteria  (maternal reported conduct disorder symptoms) |
| 189. | Piper and Corbett | 2012 | Executive function profile in the offspring of women that smoked during pregnancy | only unadjusted analyses |
| 190. | Pfinder et al. | 2012 | Explanation of social inequalities in hyperactivity/inattention in children with prenatal alcohol exposure | don't meet outcome criteria  (maternal reported ADHD symptoms) |
| 191. | Romano et al. | 2006 | Development and prediction of hyperactive symptoms from 2 to 7 years in a population-based sample | don't meet outcome criteria  (maternal reported ADHD symptoms) |
| 192. | Ruckinger et al. | 2010 | Prenatal and postnatal tobacco exposure and behavioral problems in 10-year-old children: Results from the GINI-plus prospective birth cohort study | don't meet outcome criteria  (maternal reported conduct disorder and ADHD symptoms) |
| 193. | Ruisch et al. | 2018 | Pregnancy risk factors in relation to oppositional-defiant and conduct disorder symptoms in the Avon Longitudinal Study of Parents and Children | don't meet outcome criteria  (maternal reported conduct disorder and ADHD symptoms) |
| 194. | Smidts and Oosterlaan | 2007 | How common are symptoms of ADHD in typically developing preschoolers? A study on prevalence rates and prenatal/demographic risk factors | don't meet outcome criteria  (maternal reported ADHD symptoms) |
| 195. | Van der Meer et al. | 2017 | Effects of dopaminergic genes, prenatal adversities, and their interaction on attention-deficit/hyperactivity disorder and neural correlates of response inhibition | don't meet outcome criteria  (maternal reported ADHD symptoms) |
| 196. | Motlagh et al. | 2010 | Severe psychosocial stress and heavy cigarette smoking during pregnancy: An examination of the pre- and perinatal risk factors associated with ADHD and Tourette syndrome | only unadjusted analyses |
| 197. | Monuteaux et al. | 2006 | Maternal smoking during pregnancy and offspring overt and covert conduct problems: A longitudinal study | don't meet outcome criteria  (maternal reported conduct disorder symptoms) |
| 198. | Miyake et al. | 2018 | Maternal caffeine intake in pregnancy is inversely related to childhood peer problems in Japan: The Kyushu Okinawa Maternal and Child Health Study | don't meet outcome criteria  (maternal reported conduct disorder and ADHD symptoms) |
| 199. | Milberger et al. | 1997 | Pregnancy, delivery and infancy complications and attention deficit hyperactivity disorder: Issues of gene-environment interaction | only unadjusted analyses |
| 200. | Melchior et al. | 2015 | Maternal tobacco smoking in pregnancy and children's socio-emotional development at age 5: The EDEN mother-child birth cohort study | don't meet outcome criteria  (maternal reported ADHD symptoms) |
| 201. | McGee and Stanton | 1994 | Smoking in pregnancy and child development to age 9 years | don't meet outcome criteria  (maternal reported ADHD symptoms) |
| 202. | Maughan et al. | 2001 | Pregnancy smoking and childhood conduct problems: A causal association? | don't meet outcome criteria  (maternal reported conduct disorder and ADHD symptoms) |
| 203. | Mattson et al. | 2000 | Parent ratings of behavior in children with heavy prenatal alcohol exposure and IQ-matched controls | don't meet outcome criteria  (maternal reported conduct disorder and ADHD symptoms) |
| 204. | Loomans et al. | 2012 | Caffeine intake during pregnancy and risk of problem behavior in 5- to 6-year-old children | don't meet outcome criteria  (maternal reported conduct disorder and ADHD symptoms) |
| 205. | Lavigne et al. | 2011 | Is smoking during pregnancy a risk factor for psychopathology in young children? A methodological caveat and report on preschoolers | don't meet outcome criteria  (maternal reported ADHD symptoms) |
| 206. | Linnet et al. | 2006 | Cigarette smoking during pregnancy and hyperactive-distractible preschooler's: A follow-up study | don't meet outcome criteria  (maternal reported ADHD symptoms) |
| 207. | Kukla et al. | 2008 | Maternal smoking during pregnancy, behavioral problems and school performances of their school-aged children | don't meet outcome criteria  (maternal reported measure ADHD symptoms) |
| 208. | Kovess et al. | 2015 | Maternal smoking and offspring inattention and hyperactivity: results from a cross-national European survey | don't meet outcome criteria  (maternal reported ADHD symptoms) |
| 209. | Kotimaa et al. | 2003 | Maternal smoking and hyperactivity in 8-year-old children | don't meet outcome criteria  (maternal reported ADHD symptoms) |
| 210. | Knopik et al. | 2016 | Smoking during pregnancy and ADHD risk: A genetically informed, multiple-rater approach | don't meet outcome criteria  (maternal reported ADHD symptoms) |
| 211. | Knopik et al. | 2009 | Paternal alcoholism and offspring ADHD problems: A children of twins design | don't meet outcome criteria  (maternal reported ADHD symptoms) |
| 212. | Knopik et al. | 2009 | Genetic and environmental influences on externalizing behavior and alcohol problems in adolescence: a female twin study | don't meet outcome criteria  (maternal reported conduct disorder and ADHD symptoms) |
| 213. | Knopik et al. | 2016 | Within-family effects of smoking during pregnancy on ADHD: The importance of phenotype | duplicate |
| 214. | Kieling et al. | 2013 | Gene-environment interaction in externalizing problems among adolescents: evidence from the Pelotas 1993 Birth Cohort Study | don't meet outcome criteria  (maternal reported conduct disorder and ADHD symptoms) |
| 215. | Keyes et al. | 2014 | Associations of prenatal maternal smoking with offspring hyperactivity: Causal or confounded? | don't meet outcome criteria  (maternal reported ADHD symptoms) |
| 216. | Kendler et al. | 2013 | Dimensions of Parental Alcohol Use/Problems and Offspring Temperament, Externalizing Behaviors, and Alcohol Use/Problems | don't meet outcome criteria  (maternal reported conduct disorder symptoms) |
| 217. | Kelly et al. | 2009 | Light drinking in pregnancy, a risk for behavioural problems and cognitive deficits at 3 years of age? | don't meet outcome criteria  (maternal reported conduct disorder and ADHD symptoms) |
| 218. | Kelly et al. | 2012 | Light drinking during pregnancy: Still no increased risk for socioemotional difficulties or cognitive deficits at 5 years of age? | don't meet outcome criteria  (maternal reported conduct disorder and ADHD symptoms) |
| 219. | Kahn et al. | 2003 | Role of dopamine transporter genotype and maternal prenatal smoking in childhood hyperactive-impulsive, inattentive, and oppositional behaviors | don't meet outcome criteria  (maternal reported ADHD symptoms) |
| 220. | Jaspers et al. | 2013 | Early childhood assessments of community pediatric professionals predict autism spectrum and attention deficit hyperactivity problems | don't meet outcome criteria  (maternal reported ADHD symptoms) |
| 221. | Indredavik et al. | 2007 | Prenatal smoking exposure and psychiatric symptoms in adolescence | don't meet outcome criteria  (maternal reported ADHD symptoms) |
| 222. | Mikkelsen et al. | 2017 | Maternal Caffeine Consumption during Pregnancy and Behavioral Disorders in 11-Year-Old Offspring: A Danish National Birth Cohort Study | don't meet outcome criteria  (maternal reported conduct disorder and ADHD symptoms) |
| 223. | Hutchinson et al. | 2010 | Smoking in pregnancy and disruptive behaviour in 3-year-old boys and girls: An analysis of the UK millennium cohort study | don't meet outcome criteria  (maternal reported conduct disorder and ADHD symptoms) |
| 224. | Huijbregts et al. | 2007 | Associations of maternal prenatal smoking with early childhood physical aggression, hyperactivity-impulsivity, and their co-occurrence | don't meet outcome criteria  (maternal reported ADHD symptoms) |
| 225. | Huijbregts et al. | 2008 | Hot and cool forms of inhibitory control and externalizing behavior in children of mothers who smoked during pregnancy: an exploratory study | don't meet outcome criteria  (maternal reported conduct disorder and ADHD symptoms) |
| 226. | Hannigan et al. | 2010 | A 14-year retrospective maternal report of alcohol consumption in pregnancy predicts pregnancy and teen outcomes | don't meet outcome criteria  (maternal reported conduct disorder and ADHD symptoms) |
| 227. | Han et al. | 2015 | The effects of prenatal exposure to alcohol and environmental tobacco smoke on risk for ADHD: A large population-based study | don't meet outcome criteria  (maternal reported ADHD symptoms) |
| 228. | Godel et al. | 2000 | Exposure to alcohol in utero: Influence on cognitive function and learning in a northern elementary school population | FAS sample |
| 229. | Gilman et al. | 2008 | Maternal smoking during pregnancy and children's cognitive and physical development: A causal risk factor? | don't meet outcome criteria  (observational data) |
| 230. | Gatzke-Kopp and Beauchaine | 2007 | Direct and passive prenatal nicotine exposure and the development of externalizing psychopathology | don't meet outcome criteria  (maternal reported conduct disorder and ADHD symptoms) |
| 231. | Fried et al. | 1992 | A follow-up study of attentional behavior in 6-year-old children exposed prenatally to marijuana, cigarettes, and alcohol | don't meet outcome criteria  (maternal reported ADHD symptoms) |
| 232. | Fitzpatrick et al. | 2014 | Parental bad habits breed bad behaviors in youth: Exposure to gestational smoke and child impulsivity | don't meet outcome criteria  (teacher reported ADHD symptoms) |
| 233. | Fergusson et al. | 1993 | Maternal smoking before and after pregnancy: Effects on behavioral outcomes in middle childhood | don't meet outcome criteria  (maternal reported conduct disorder and ADHD symptoms) |
| 234. | Estabrook et al. | 2016 | Separating Family-Level and Direct Exposure Effects of Smoking During Pregnancy on Offspring Externalizing Symptoms: Bridging the Behavior Genetic and Behavior Teratologic Divide | don't meet outcome criteria  (maternal reported conduct disorder, ODD and ADHD symptoms) |
| 235. | Ellingson et al. | 2014 | A sibling-comparison study of smoking during pregnancy and childhood psychological traits | don't meet outcome criteria  (maternal reported conduct disorder, ODD and ADHD symptoms) |
| 236. | Eiden et al. | 2018 | Pre- and postnatal tobacco and cannabis exposure and child behavior problems: Bidirectional associations, joint effects, and sex differences | don't meet outcome criteria  (maternal reported ADHD symptoms) |
| 237. | Eichler et al. | 2018 | Effects of prenatal alcohol consumption on cognitive development and ADHD-related behaviour in primary-school age: a multilevel study based on meconium ethyl glucuronide | don't meet outcome criteria  (maternal reported ADHD symptoms) |
| 238. | Durr et al. | 2015 | Tobacco smoking during pregnancy and risk of adverse behaviour in offspring: A follow-up study | don't meet outcome criteria  (maternal reported conduct disorder and ADHD symptoms) |
| 239. | Dolan et al. | 2016 | Testing Causal Effects of Maternal Smoking During Pregnancy on Offspring's Externalizing and Internalizing Behavior | don't meet outcome criteria  (maternal reported ADHD symptoms) |
| 240. | Dodge et al. | 2014 | Protective effects of the alcohol dehydrogenase-ADH1B*3 allele on attention and behavior problems in adolescents exposed to alcohol during pregnancy | don't meet outcome criteria  (teacher reported conduct disorder, ODD and ADHD symptoms) |
| 241. | Disney et al. | 2008 | Strengthening the case: prenatal alcohol exposure is associated with increased risk for conduct disorder | don't meet outcome criteria  (maternal reported conduct disorder symptoms) |
| 242. | Desrosiers et al. | 2013 | Associations between prenatal cigarette smoke exposure and externalized behaviors at school age among Inuit children exposed to environmental contaminants | don't meet outcome criteria  (teacher reported conduct disorder, ODD and ADHD symptoms) |
| 243. | Sutin et al. | 2017 | Maternal cigarette smoking during pregnancy and the trajectory of externalizing and internalizing symptoms across childhood: Similarities and differences across parent, teacher, and self reports | don't meet outcome criteria  (maternal reported conduct disorder and ADHD symptoms) |
| 244. | Sood et al. | 2001 | Prenatal alcohol exposure and childhood behavior at age 6 to 7 years: I. dose-response effect | don't meet outcome criteria  (maternal reported ADHD symptoms) |
| 245. | Silberg et al. | 2003 | Maternal smoking during pregnancy and risk to boys' conduct disturbance: An examination of the causal hypothesis | don't meet outcome criteria  (child's reported conduct disorder symptoms) |
| 246. | Sen and Swaminathan | 2007 | Maternal prenatal substance use and behavior problems among children in the U.S | don't meet outcome criteria  (maternal reported ADHD symptoms) |
| 247. | Day et al. | 2013 | The association between prenatal alcohol exposure and behavior at 22 years of age | don't meet outcome criteria  (maternal reported ADHD symptoms) |
| 248. | Day et al. | 2000 | Effects of prenatal tobacco exposure on preschoolers' behavior | don't meet outcome criteria  (maternal reported ODD and ADHD symptoms) |
| 249. | D'Onofrio et al. | 2008 | Smoking during pregnancy and offspring externalizing problems: An exploration of genetic and environmental confounds | don't meet outcome criteria  (maternal reported conduct disorder, ODD and ADHD symptoms) |
| 250. | D'Onofrio et al. | 2007 | Causal inferences regarding prenatal alcohol exposure and childhood externalizing problems | don't meet outcome criteria  (maternal reported conduct disorder and ADHD symptoms) |
| 251. | Cornelius et al. | 2007 | Smoking during teenage pregnancies: Effects on behavioral problems in offspring | don't meet outcome criteria  (maternal reported ADHD symptoms) |
| 252. | Cornelius et al. | 2012 | Long-term effects of prenatal cigarette smoke exposure on behavior dysregulation among 14-year-old offspring of teenage mothers | don't meet outcome criteria  (maternal reported ADHD symptoms) |
| 253. | Cornelius et al. | 2012 | Prenatal cigarette smoking: Long-term effects on young adult behavior problems and smoking behavior | externalising disorder not specified |
| 254. | Cornelius et al. | 2011 | Effects of prenatal cigarette smoke exposure on neurobehavioral outcomes in 10-year-old children of adolescent mothers | don't meet outcome criteria  (maternal reported ADHD symptoms) |
| 255. | Clark et al. | 2016 | Developmental pathways from prenatal tobacco and stress exposure to behavioral disinhibition | don't meet outcome criteria  (maternal reported ADHD symptoms) |
| 256. | Chiu et al. | 2009 | Demographic and perinatal factors for behavioral problems among children aged 4-9 in Taiwan: Regular article | don't meet outcome criteria  (maternal reported conduct disorder and ADHD symptoms) |
| 257. | Chiodo et al. | 2010 | The Impact of Maternal Age on the Effects of Prenatal Alcohol Exposure on Attention | don't meet outcome criteria  (teacher reported ADHD symptoms) |
| 258. | Carter et al. | 2008 | Maternal smoking during pregnancy and behaviour problems in a birth cohort of 2-year-old Pacific children in New Zealand | don't meet outcome criteria  (maternal reported ADHD symptoms) |
| 259. | Button et al. | 2005 | Relationship between antisocial behaviour, attention-deficit hyperactivity disorder and maternal prenatal smoking | don't meet outcome criteria  (maternal reported ADHD symptoms) |
| 260. | Buschgens et al. | 2009 | Externalizing behaviors in preadolescents: Familial risk to externalizing behaviors, prenatal and perinatal risks, and their interactions | don't meet outcome criteria  (maternal reported conduct disorder and ADHD symptoms) |
| 261. | Brown et al. | 1991 | Effects of prenatal alcohol exposure at school age. II. Attention and behavior | don't meet outcome criteria  (maternal and teacher reported conduct disorder and ADHD symptoms) |
| 262. | Brion et al. | 2010 | Maternal smoking and child psychological problems: Disentangling causal and noncausal effects | don't meet outcome criteria  (maternal reported conduct disorder and ADHD symptoms) |
| 263. | Brinksma et al. | 2017 | Age-dependent role of pre- and perinatal factors in interaction with genes on ADHD symptoms across adolescence | don't meet outcome criteria  (maternal reported ADHD symptoms) |
| 264. | Brennan et al. | 2011 | Interactions between the COMT Val108/158Met polymorphism and maternal prenatal smoking predict aggressive behavior outcomes | don't meet outcome criteria  (child's, maternal and teacher reported ADHD symptoms) |
| 265. | Bos-Veneman et al. | 2010 | Role of perinatal adversities on tic severity and symptoms of attention deficit/hyperactivity disorder in children and adolescents with a tic disorder | don't meet outcome criteria  (maternal reported ADHD symptoms) |
| 266. | Boden et al. | 2010 | Risk factors for conduct disorder and oppositional/defiant disorder: Evidence from a New Zealand birth cohort | don't meet outcome criteria  (maternal reported ODD and ADHD symptoms) |
| 267. | Bekkhus et al. | 2010 | Intrauterine exposure to caffeine and inattention/overactivity in children | don't meet outcome criteria  (maternal reported ADHD symptoms) |
| 268. | Becker et al. | 2008 | Interaction of Dopamine Transporter Genotype with Prenatal Smoke Exposure on ADHD Symptoms | don't meet outcome criteria  (maternal reported ADHD symptoms) |
| 269. | Batstra et al. | 2003 | Effect of antenatal exposure to maternal smoking on behavioural problems and academic achievement in childhood: Prospective evidence from a Dutch birth cohort | don't meet outcome criteria  (maternal reported ADHD symptoms) |
| 270. | Arruda et al. | 2015 | ADHD and mental health status in Brazilian school-age children | only unadjusted analyses |
| 271. | Anselmi et al. | 2010 | Early determinants of attention and hyperactivity problems in adolescents: the 11-year follow-up of the 1993 Pelotas (Brazil) birth cohort study | externalising disorder not specified |
| 272. | Agrawal et al. | 2010 | The effects of maternal smoking during pregnancy on offspring outcomes | don't meet outcome criteria  (maternal reported conduct disorder and ADHD symptoms) |
| 273. | Sayal et al. | 2007 | Prenatal alcohol exposure and gender differences in childhood mental health problems: A longitudinal population-based study | don't meet outcome criteria  (maternal and teacher reported conduct disorder and ADHD symptoms) |
| 274. | Sayal et al. | 2009 | Binge pattern of alcohol consumption during pregnancy and childhood mental health outcomes: longitudinal population-based study | don't meet outcome criteria  (maternal reported conduct disorder and ADHD symptoms) |
| 275. | Sayal et al. | 2014 | Prenatal exposure to binge pattern of alcohol consumption: mental health and learning outcomes at age 11 | don't meet outcome criteria  (maternal and teacher reported conduct disorder and ADHD symptoms) |
| 276. | Salationo-Oliveira et al. | 2016 | COMT and prenatal maternal smoking in associations with conduct problems and crime: the Pelotas 1993 birth cohort study | don't meet outcome criteria  (maternal reported conduct disorder symptoms) |
| 277. | Rodriguez et al. | 2009 | Is prenatal alcohol exposure related to inattention and hyperactivity symptoms in children? Disentangling the effects of social adversity | don't meet outcome criteria  (maternal and teacher reported ADHD symptoms) |
| 278. | Rodriguez and Bohlin | 2005 | Are maternal smoking and stress during pregnancy related to ADHD symptoms in children? | don't meet outcome criteria  (maternal reported ADHD symptoms) |
| 279. | Pourcain et al. | 2011 | Links between co-occurring social-communication and hyperactive-inattentive trait trajectories | don't meet outcome criteria  (maternal reported ADHD symptoms) |
| 280. | Piper et al. | 2012 | Maternal smoking cessation and reduced academic and behavioral problems in offspring | don't meet outcome criteria  (maternal reported ADHD symptoms) |
| 281. | Pfinder et al. | 2014 | Impact of moderate prenatal alcohol exposure on problem behaviors in preschool and school children stronger effects in disadvantaged populations? Results from the KiGGS study | don't meet outcome criteria  (maternal reported conduct disorder and ADHD symptoms) |
| 282. | Parker et al. | 2016 | Prenatal smoking and childhood behavior problems: is the association mediated by birth weight? | don't meet outcome criteria  (maternal reported conduct disorder and ADHD symptoms) |
| 283. | Park et al. | 2014 | Mediating role of stress reactivity in the effects of prenatal tobacco exposure on childhood mental health outcomes | don't meet outcome criteria  (maternal and teacher reported conduct disorder and ADHD symptoms) |
| 284. | Palmer et al. | 2016 | Effects of Maternal Smoking during Pregnancy on Offspring Externalizing Problems: Contextual Effects in a Sample of Female Twins | don't meet outcome criteria  (child's and maternal reported conduct disorder and ADHD symptoms) |
| 285. | Palili et al. | 2011 | Inattention, hyperactivity, impulsivity--epidemiology and correlations: a nationwide greek study from birth to 18 years | don't meet outcome criteria  (maternal reported ADHD symptoms) |
| 286. | Orlebeke et al. | 1999 | Child behavior problems increased by maternal smoking during pregnancy | don't meet outcome criteria  (maternal reported conduct disorder and ADHD symptoms) |
| 287. | Orlebeke et al. | 1997 | Increase in child behavior problems resulting from maternal smoking during pregnancy | don't meet outcome criteria  (maternal reported conduct disorder and ADHD symptoms) |
| 288. | Obel et al. | 2009 | Smoking during pregnancy and hyperactivity-inattention in the offspring - Comparing results from three Nordic cohorts | don't meet outcome criteria  (maternal reported ADHD symptoms) |
| 289. | O'Leary et al. | 2010 | Evidence of a complex association between dose, pattern and timing of prenatal alcohol exposure and child behaviour problems | don't meet outcome criteria  (maternal reported conduct disorder and ADHD symptoms) |
| 290. | Noordermeer et al. | 2017 | Risk factors for comorbid oppositional defiant disorder in attention-deficit/hyperactivity disorder | no exposure-outcome association measured |
| 291. | Niclasen et al. | 2014 | Prenatal exposure to alcohol, and gender differences on child mental health at age seven year | don't meet outcome criteria  (maternal reported conduct disorder and ADHD symptoms) |
| 292. | Niclasen et al. | 2014 | Is alcohol binge drinking in early and late pregnancy associated with behavioural and emotional development at age 7 years? | don't meet outcome criteria  (maternal reported conduct disorder and ADHD symptoms) |
| 293. | Murray et al. | 2010 | Very early predictors of conduct problems and crime: results from a national cohort study | don't meet outcome criteria  (maternal reported conduct disorder symptoms) |
| 294. | Murray et al. | 2016 | Moderate alcohol drinking in pregnancy increases risk for children's persistent conduct problems: causal effects in a Mendelian randomisation study | don't meet outcome criteria  (maternal reported conduct disorder symptoms) |
| 295. | Hill et al. | 2000 | Maternal smoking and drinking during pregnancy and the risk for child and adolescent psychiatric disorders” - did not any adjusted results on tobacco and alcohol exposure and ADHD, CD and ODD | results not reported |
| 296. | Tzoumakis et al. | 2018 | Prenatal maternal smoking, maternal offending, and offspring behavioural and cognitive outcomes in early childhood | don't meet outcome criteria (maternal reported ADHD symptoms) |
| 297. | Vergunst et al. | 2019 | Multi-rater developmental trajectories of hyperactivity-impulsivity and inattention symptoms from 1.5 to 17years: a population-based birth cohort study | don't meet outcome criteria (maternal and teacher reported ADHD symptoms) |
| 298. | Sellers et al. | 2020 | Using a cross-cohort comparison design to test the role of maternal smoking in pregnancy in child mental health and learning: evidence from two UK cohorts born four decades apart | don't meet outcome criteria (maternal reported ADHD and CD symptoms) |
| 299. | Minatoya et al. | 2019 | Prenatal tobacco exposure and ADHD symptoms at pre-school age: the Hokkaido Study on Environment and Children's Health | don't meet outcome criteria (maternal reported ADHD symptoms) |
| 300. | MaMaison et al. | 2018 | Prevalence and risk factors of psychiatric disorders in early adolescence: 2004 Pelotas (Brazil) birth cohort | no exposure-outcome association measured |
| 301. | Doyle et al. | 2019 | Relation Between Oppositional/Conduct Behaviors and Executive Function Among Youth with Histories of Heavy Prenatal Alcohol Exposure | FAS sample |
| 302. | Gutman et al. | 2019 | Developmental Trajectories of Conduct Problems and Cumulative Risk from Early Childhood to Adolescence | don't meet outcome criteria (maternal reported CD symptoms) |
| 303. | Hartman and Craig | 2018 | Examining the Association Between Maternal Smoking During Pregnancy and Child Behavior Problems Using Quality-Adjusted Life Years | externalising disorder not specified |
| 304. | Dong et al. | 2018 | Prenatal exposure to maternal smoking during pregnancy and attention-deficit/hyperactivity disorder in offspring: A meta-analysis | review/meta-analysis |
| 305. | Driga and Drigas | 2019 | ADHD in the Early Years: Pre-Natal and Early Causes and Alternative Ways of Dealing | wrong study design |
| 306. | Cluver et al. | 2019 | The association of prenatal alcohol exposure on the cognitive abilities and behaviour profiles of 4-year-old children: a prospective cohort study | don't meet outcome criteria (maternal reported ADHD symptoms) |
| 307. | Cao et al. | 2020 | Early tobacco smoke exposure, preschool cool/hot inhibitory control, and young adolescents' externalizing/internalizing problems | externalising disorder not specified |
| 308. | Brannigan et al. | 2020 | Prenatal tobacco exposure and psychiatric outcomes in adolescence: is the effect mediated through birth weight? | externalising disorder not specified |
| 309. | Baykal et al. | 2018 | Prenatal and postnatal characteristics in attention deficit hyperactivity disorder | Not English language |
| 310. | Easey et al. | 2019 | Prenatal alcohol exposure and offspring mental health: A systematic review | review/meta-analysis |
| 311. | Ekblad et al. | 2020 | The Effect of Smoking during Pregnancy on Severity and Directionality of Externalizing and Internalizing Symptoms: A Genetically Informed Approach | externalising disorder not specified |
| 312. | He et al. | 2020 | Maternal Smoking During Pregnancy and ADHD: Results From a Systematic Review and Meta-Analysis of Prospective Cohort Studies | review/meta-analysis |
| 313. | Koch et al. | 2020 | Mental disorders in referred 0-3-year-old children: a population-based study of incidence, comorbidity and perinatal risk factors | no exposure-outcome association measured |
| 314. | Kim et al. | 2020 | Environmental risk factors, protective factors, and peripheral biomarkers for ADHD: an umbrella review | review/meta-analysis |
| 315. | Koponen et al. | 2020 | Prenatal substance exposure, adverse childhood experiences and diagnosed mental and behavioral disorders - A longitudinal register-based matched cohort study in Finland | externalising disorder not specified |
| 316. | Massey et al. | 2020 | Dimension- and context-specific expression of preschoolers' disruptive behaviors associated with prenatal tobacco exposure | wrong outcomes |
| 317. | Nissinen et al. | 2021 | Completed secondary education among youth with prenatal substance exposure: A longitudinal register-based matched cohort study | externalising disorder not specified |
| 318. | Porter et al. | 2019 | Low-moderate prenatal alcohol exposure and offspring attention-deficit hyperactivity disorder (ADHD): systematic review and meta-analysis | review/meta-analysis |
| 319. | Rolan et al. | 2019 | Contextual Influences of prenatal environments on executive function risk for adolescent substance use and externalizing behavior | conference/meeting abstract |
| 320. | Ruisch et al. | 2019 | Interplay between genome-wide implicated genetic variants and environmental factors related to childhood antisocial behavior in the UK ALSPAC cohort | no comparison/control group |
| 321. | Sandtorv et al. | 2018 | Symptoms Associated With Attention Deficit/Hyperactivity Disorder and Autism Spectrum Disorders in School-Aged Children Prenatally Exposed to Substances | wrong exposure |
| 322. | Schlack et al. | 2018 | Predictors of Stability of Parent Report on ADHD Lifetime Prevalence and Incidence of Parent-Reported ADHD Diagnosis in the Developmental Course Over Six Years - Results from the KiGGS Study | Not English language |
| 323. | Sengupta et al. | 2020 | Facing the Methodological Challenge in Dissecting the Genetics of ADHD: A Case for Deep Phenotyping and Heterogeneity Reduction | no comparison/control group |
| 324. | Tole et al. | 2019 | The role of pre-, peri-, and postnatal risk factors in bipolar disorder and adult ADHD | only unadjusted results |
| 325. | Waltes et al. | 2019 | Impact of autism-associated genetic variants in interaction with environmental factors on ADHD comorbidities: an exploratory pilot study | no comparison/control group |
| 326. | Zambrano-Sanchez et al. | 2021 | Maternal smoking during pregnancy and physiological anxiety in children with attention deficit hyperactivity disorder | wrong outcomes |
| 327. | Michelle et al. | 2020 | Prenatal alcohol exposure and risk of attention deficit hyperactivity disorder in offspring: A retrospective analysis of the millennium cohort study | duplicate |
| 328. | Ekblad et al. | 2020 | The Effect of Smoking during Pregnancy on Severity and Directionality of Externalizing and Internalizing Symptoms: A Genetically Informed Approach | externalising disorder not specified |
| 329. | Marceau et al. | 2019 | Parenting and Prenatal Risk as Moderators of Genetic Influences on Conduct Problems During Middle Childhood | don't meet outcome criteria  (maternal reported conduct disorder symptoms) |
| 330. | Ichikawa et al. | 2018 | Prenatal Alcohol Exposure and Child Psychosocial Behavior: A Sibling Fixed-Effects Analysis | don't meet outcome criteria  (maternal reported ADHD symptoms) |
| 331. | Lund et al. | 2019 | Is the association between maternal alcohol consumption in pregnancy and pre-school child behavioural and emotional problems causal? Multiple approaches for controlling unmeasured confounding | don't meet outcome criteria  (maternal reported ADHD symptoms) |

### Supplementary Table S4. Studies not reporting all the results

| **Author/Year** | **Title** | **Country** | **Study design** | **Exposure** | **Outcome** | **Missing data** |
| --- | --- | --- | --- | --- | --- | --- |
| Talati et al., 2016 | Brain derived neurotrophic factor moderates associations between maternal smoking during pregnancy and offspring behavioral disorders” | USA | Longitudinal family study | Smoking | ADHD  CD | Results not reported on ADHD due to the low number of cases |
| Talati et al., 2017 | Prenatal tobacco exposure, birthweight, and offspring psychopathology” – did not report ADHD results due to the low number of cases | USA | Longitudinal family study | Smoking | ADHD  CD | Results not reported on ADHD due to the low number of cases |
| Whitbeck et al., 2009 | Gestational risks and psychiatric disorders among indigenous adolescents” – did not report results on tobacco exposure and ADHD | USA | Cohort study | Smoking  Alcohol | ADHD  CD | Results on tobacco exposure not reported |
| Hill et al., 2000 | Maternal smoking and drinking during pregnancy and the risk for child and adolescent psychiatric disorders” - did not any adjusted results on tobacco and alcohol exposure and ADHD, CD and ODD | USA | Longitudinal family study | Smoking  Alcohol | ADHD  CD  ODD | Adjusted results not reported |
| Kim et al., 2009 | Perinatal and Familial Risk Factors Are Associated with Full Syndrome and Subthreshold Attention-Deficit Hyperactivity Disorder in a Korean Community Sample | Korea | Case-control | Smoking  Alcohol  Caffeine | ADHD | Results not reported on tobacco exposure due to the low prevalence of smoking in the sample |
| Knopik et al., 2006 | Maternal alcohol use disorder and offspring ADHD: disentangling genetic and environmental effects using a children-of-twins design | Australia | Longitudinal twin study | Smoking  Alcohol | ADHD | Only unadjusted results reported for alcohol exposure |

### Supplementary Table S5. Study characteristics and results of cohort, longitudinal and cross-sectional studies

| **Author/Year** | **Country** | **Study sample** | **Cases/total sample size** | **Gender** | **Offspring age at assessment** | **Exposure assessment** | **Classification of exposure** | **Outcome** | **Outcome assessment** | **Follow up time** | **Confounders** | **Results (as reported in the study)** |
| --- | --- | --- | --- | --- | --- | --- | --- | --- | --- | --- | --- | --- |
| **Smoking** |  |  |  |  |  |  |  |  |  |  |  |  |
| Ball et al., 2010 | USA | Prospective longitudinal cohort study in general population (the Collaborative Perinatal Project (CPP), the new England Family Study (NEFS) and Providence cohort | 219/2024 | Both | 37.2 years | Prospective self-report during all the pregnancy trimesters | No smoking (ref); <half pack per day; >half pack to <full pack; <full pack | ADHD | Retrospective assessment using Diagnostic interview Schedule (DIS) | 1, 4, 7 years and adulthood | Ascertainment source, family psychopathology, maternal education, offspring gender | **OR (95% CI)**  No smoking (ref)  <half pack per day: 1.2 (0.81, 1.7)  >halff pack to <full pack: 1.1 (0.75, 1.6)  >full pack: 1.1 (0.67, 1.8)" |
| Gustavson et al., 2017* | Norway | Prospective longitudinal birth cohort study in general population (the Norwegian Mother, Father and Child Cohort study (MoBa)) | 2035/104,846 | Both | 5 years | Prospective self-report during the 2nd and 3rd pregnancy trimester | Binary (Yes/No) | ADHD | ADHD diagnosis obtained from the Norwegian Patient Registry (NPR) | 6, 18 months, 3, 5, 7, 8 and 13 years (ongoing study) | Parental age, education, parental ADHD symptoms; maternal (pre-pregnancy) and paternal BMI; maternal alcohol consumption during pregnancy; maternal parity; child’s birth year; geographical residential region | **Hazard ratio (HR) (95% CI), p-value**  Maternal smoking:  1.48 (1.3, 1.68), <0.001  Paternal smoking:  1.28 (1.16, 1.42)  **the magnitude of the effect size was similar in all negative controls (paternal smoking, grandmother's smoking when pregnant with the mother, and maternal smoking in previous pregnancies)* |
| Langley et al., 2012 | UK | Prospective longitudinal birth cohort study in general population (the Avon Longitudinal Study of Parents and Children (ALSPAC)) | 121/5637 | Both | 7.6 years | Prospective self-report during the 2nd and 3rd pregnancy trimester | No smoking (ref);  1-9 cigarettes/day; 10-19 cigarettes/day;  >20 cigarettes/day | ADHD | the Development and Well-Being Assessment (DAWBA) and diagnosis were generated based on maternal and teacher reports | From birth to adulthood (ongoing study) | child’s sex, ethnicity, multiple births (twins), maternal prenatal alcohol use, social class | **OR (95% CI), p-value**  1.72 (1.14, 2.61), <0.05  **the magnitude of the effect estimates were similar with paternal smoking* |
| Obel et al., 2011 | Finland | Population based cohort and registry study | 7023/868,449 6013 boys 1010 girls | Both (reported separately for boys and girls) | 15 years | Information collected during the routine visit to midwives in the 1st and 2nd pregnancy trimester | Non-smokers (ref); Smokers  Non-smokers (ref);  1-9 cigarettes/day 10+ cigarettes/day Non-smokers (ref); Only 1st trimester; After first trimester | Hyperkinetic disorder | the Finnish Hospital Discharge Register (FHDR) | 15 years | child age, birth year, child sex, gestational age at birth, parity, maternal age, SES | **HR (95% CI), p-value**  Total sample: 2.01 (1.9, 2.12)  Boys: 1.96 (1.85, 2.08)  Girls: 2.28 (1.99, 2.63)  Total sample:  Overall:  1.92 (1.69, 2.18)  1-9 cigarettes:  1.77 (1.47, 2.12)  10+ cigarettes:  2.03 (1.74, 2.37)  Boys:  Overall: 1.88 (1.64, 2.15)  1-9 cigarettes: 1.75 (1.43, 2.13)  10+ cigarettes: 1.97 (1.67, 2.33)  Girls:  Overall: 2.23 (1.56, 3.18)  1-9 cigarettes: 1.93 (1.15, 3.23)  10+ cigarettes: 2.46 (1.59, 3.81)  Total sample:  Overall: 1.92 (1.80, 2.04) Only 1st trimester: 1.34 (1.11, 1.62)  After 1st trimester: 1.98 (1.86, 2.12)  Boys:  Overall: 1.88 (1.75, 2.01) Only 1st trimester:  1.34 (1.09, 1.64)  After 1st trimester:  1.94 (1.81, 2.08)  Girls:  Overall: 2.13 (1.82, 2.49) Only 1st trimester:  1.35 (0.82, 2.22)  After 1st trimester: 2.21 (1.88, 2.60)  **no difference in effect estimates in sibling comparison analyses* |
| Obel et al., 2016* | Denmark | Population based cohort and registry study | 17,381/968,665 13,524 boys 3857 girls | Both (reported separately for boys and girls) | 9 years | Information collected during the visits to general practitioners in each pregnancy trimester | Non-smokers (ref); Smokers  Non-smokers (ref); Quitted smoking during 1st trimester; Continued smoking Non-smokers (ref); 1-10cigarettes/day; 10+ cigarettes | Hyperkinetic disorder | ADHD diagnosis obtained from the ICD-10 registration system and ADHD medication obtained from the Register of Medicinal Product Statistics | Median follow up time 9 years | birth year, child sex, maternal age, and parity | **HR (95% CI)**  Total sample:  2.01 (1.94, 2.07)  Boys: 1.98 (1.91, 2.05)  Girls: 2.12 (1.99, 2.27)  No smoking (ref)  Quitted smoking during 1st trimester:  1.61 (1.39, 1.88)  Continued smoking:  2.19 (2.09, 2.29)  No smoking (ref)  1-10 cigarettes:  1.92 (1.81, 2.02)  10+ cigarettes:  2.77 (2.60, 2.95)  **no associations in sibling analyses*  1-10 cigarettes:  1.17 (0.86, 1.6)  10+ cigarettes:  1.08 (0.72, 1.61) |
| Skoglund et al., 2014* | Sweden | Population based registry study | 19,891/768,227 | Both | ~9 years | Antenatal visit during the 1st pregnancy trimester | No smoking (ref); Moderate (1-9 cigarettes);  High (>10 cigarettes) | ADHD | Diagnosis and medication use of ADHD were identified from the Patient Register and the Swedish Prescribed Drug register | Up to 9 years | child sex, birth year, parity, maternal age at child’s birth, cohabitation with the child's father, maternal education, mother's country of birth | **HR (95% CI)**  Moderate (1-9 cigarettes): 1.62 (1.56, 1.69)  High (>10 cigarettes):  2.04 (1.95, 2.13)  **no association in sibling and cousin analyses*  Moderate (1-9 cigarettes): 0.88 (0.73, 1.06)  High (>10 cigarettes):  0.84 (0.65, 1.06) |
| Zhu et al., 2014* | Denmark | Population based longitudinal and registry study (Danish Birth Cohort Study) | 2009/84,803 | Both | >5 years | Interview during the 2nd pregnancy trimester | Non-smoker (ref); Smoker;  Nicotine replacement user; Smoking quitter | ADHD | A combination of ADHD medication and hospital diagnosis (the International Classification of Diseases, 10th Revision [ICD-10]) was used. | Up to 8 to 14 years | maternal age at birth of the child, alcohol intake during pregnancy, parental socio-occupational status, parental psychopathology, parity and child’s gender | **HR (95% CI)**  Smoking (both parents): 1.83 (1.60, 2.10)  Smoking (father non-smoker): 1.63 (1.36, 1.94)  Paternal smoking (mother non-smoker)  1.29 (1.14, 1.47)  **stronger association with maternal smoking and nicotine replacement when father was non-smoker* |
| Lindblad et al., 2010* | Sweden | Population based registry study | 6141/927,007 | Both | 6-19 years | Visit to the midwives during the 1st pregnancy trimester | No smoking in any pregnancy (ref);  No smoking at least in one pregnancy;  1-9 cigarettes/day; 10+ cigarettes/day | ADHD | ADHD medication use was obtained from The Swedish Prescribed Drug Register | from 6 up to 19 years | maternal age and year of birth, child sex, and parity. Marital status, maternal education, social assistance, parental psychiatric disorders and substance use, child’s indicator of small for gestational age (gestational age and Apgar score) | **OR (95% CI)**  No smoking at least in one pregnancy: 1.43 (1.26, 1.61) 1-9 cigarettes: 1.59 (1.49, 1.70)  10+ cigarettes: 1.89 (1.75, 2.04)  **no association within-mother between pregnancy analyses*  1-9 cigarettes: 0.96 (0.73, 1.13)  10+ cigarettes: 1.26 (0.95, 1.58) |
| Braun et al.., 2006 | USA | population based cross-sectional study (The National Health and Nutrition Examination Survey (NHANES)) | 135/4704 | Both (reported separately for boys and girls) | 4-15 years | Not specified | Binary (Yes/No) | ADHD | Combination of parent report whether child has diagnosed for ADHD and stimulant medication use based on National Drug Codes | NA | child’s age, sex, race, socioeconomic status as measured by poverty-to-income ratio (PIR), mother’s age at child’s birth, national insurance coverage, preschool attendance and perinatal distress (birth weight and admission to neonatal intensive care unit) | **OR (95% CI), p-value**  Total sample: 2.5 (1.2, 5.2), 0.02  Boys: 2.1 (0.9, 4.7), 0.07 Girls: 4.6 (1.7, 12.4) |
| Wakschlag et al., 1997 | USA | longitudinal study (clinic referred sample) | 105/177 | only boys | 12-17 years | retrospective self-report (pregnancy trimester not specified) | No and occasional smoking (ref); Smoked <half pack/day;  Smoked >half pack/day | CD | A diagnosis was derived by combining parent, teacher and child report of the National Institute of Mental Health Diagnostic Interview Schedule for Children (DISC) | 4 annual assessments (starting age 7 years) and 5th assessment in 2 years | SES and ethnicity, parental psychopathologic conditions (paternal antisocial personality disorder, maternal MMPI antisocial index, and maternal and paternal depression and substance abuse), other pregnancy risk factors (maternal age at child’s birth, use of illicit drugs and alcohol during pregnancy, pregnancy and birth complications, low birth weight, and prematurity), and family and parenting risk factors (number of children in the household, marital status, poor communication, little supervision, and ineffective and harsh discipline | **OR (95% CI), p-value** Smoked <half pack/day:  1.6 (0.53, 5.11), 0.39  Smoked >half pack/day:  3.3 (1.2, 9.07), 0.02 |
| Talati et al., 2016 | USA | longitudinal family study (clinical sample of depressed probands) | - /209 | both | not specified | retrospective self-report (pregnancy trimester not specified) | Binary (smoked >10 cigarettes/did not smoke >10 cigarettes/day) | CD | Semi-structured interview (Schedule for Affective Disorders and Schizophrenia, SADS) administered by trained mental health professional | Six waves up to 30 years | age, gender, and maternal depression and substance use history | Effect estimate (SE), p-value Smoking: 0.15 (0.51), 0.76 Smoking and child genotype interaction:  1.92 (0.81), 0.017 |
| Braun et al., 2008 | USA | population based cross-sectional study (The National Health and Nutrition Examination Survey (NHANES)) | 68/3081 | both | 8-15 years | retrospective self-report (pregnancy trimester not specified) | Binary (Yes/No) | CD | the Diagnostic Interview Schedule for Children (DISC) based on DSM-IV criteria | NA | child’s age, sex, race, socioeconomic status as measured by poverty-to-income ratio (PIR), and mother’s age at child’s birth | **OR (95% CI)**  3.00 (1.36, 6.63) |
| Ellis et al., 2012 | Norway | population based prospective cohort study (the Trondheim Early Secure Study (TESS)) | 995 34 ADHD cases 57 ODD cases | both | 4 years | retrospective interview (assessed in each pregnancy trimester) | Binary (Yes/No) | ADHD  ODD | Structured interview with mothers using the Preschool Age Psychiatric Assessment (PAPA - diagnosis based on DSM-IV criteria) | 4 years (ongoing) | Personality traits of narcissistic, histrionic, borderline, schizotypal, paranoid, avoidant, dependent, obsessive-compulsive personality, parental alcohol use, parental anxiety, depression, prenatal stress, depression, alcohol use, planned pregnancy, parental feelings about pregnancy, mothers’ feelings in the first month after birth, parental experience of mental breakdown, parent requested medical treatment, parent ever been arrested, parent ever been indicted by police, parental ability to pay family expenses, parent received medical treatment for psychological disorder, and parental admission to a mental health institution. Maternal age and SES | **OR (95% CI), p-value**  ADHD: 2.17 (1.3, 3.61), 0.003  ODD: 2.46 (1.66, 3.63), 0.001  **propensity score analyses* |
| Nigg et al., 2007 | USA | longitudinal cohort study | 798 95  ADHD cases  46 CD cases  89 ODD cases | both | 11 years in ADHD  17 years in CD and ODD | retrospective self-report (pregnancy trimester not specified) | Binary (Yes/No) | ADHD  CD  ODD | the Diagnostic Interview Schedule for Children (DISC; version 2.1) | 3 assessments: at age 6, 11 and 17 years | low birth weight, maternal substance use disorders, maternal education, home environment (urban vs suburban) | **OR (95% CI), p-value**  ADHD: 1.19 (0.79, 1.8)  CD: 2.32 (1.05, 5.15), <0.05 ODD: 2.31 (1.24, 4.31), <0.05  **low birth weight did not mediate the association with ADHD. Independent effect of low birth weight was observed on ADHD but not on CD or ODD* |
| Talati et al., 2017 | USA | longitudinal family study (clinical sample of depressed mothers) | 238  21 ADHD cases 66 CD cases | Both (reported separately for boys and girls) | 6-17 years | retrospective self-report (pregnancy trimester not specified) | Binary (whether or not mother smoked >10 cigarettes/day) | CD | Semi-structured interview (Schedule for Affective Disorders and Schizophrenia, SADS) administered by trained mental health professional | 6 waves (age 1, 2, 10, 20 and 30 years). Mean follow-up time 27.7 years | Familial risk for depression, offspring age at final interview, and sex (except in the sex-specific models). Further covariates included lifetime diagnosis of nicotine dependence in either parent and any parental mental health diagnosis | **OR (95% CI), p-value**  Total sample:  1.75 (0.94, 3.23), <0.07  Boys: 2.63 (1.05, 6.49), <0.05  Girls: 1.08 (0.41, 2.82)  **birth weight did not mediate the association* |
| Weissman et al., 1999 | USA | longitudinal family study (clinical sample of depressed mothers) | -/147 | Both (reported separately for boys and girls) | 16.4 years | retrospective self-report (pregnancy trimester not specified) | Binary (no smoking; 10+ cigarettes/day) | CD  ADHD | Parent and offspring interview using the Schedule for Affective Disorders and Schizophrenia-Lifetime version (SADS-LA) | 3 assessments in 10 years: baseline 6 to 23 years | history of maternal depression, offspring age and divorce | **Relative Risk (RR) (95% CI)** CD 13-17 years:  Boys: 1.70 (0.48, 5.96)  Girls: 1.00 (0.35, 2.86) ADHD <13 years:  Boys: 0.44 (0.094, 2.09) Girls: 2.16 (0.135, 34.71) |
| Nomura et al., 2010 | USA | longitudinal study | 209 65 ADHD cases 6 ODD cases | both | 4.3 years | retrospective self-report | Binary (Yes/No) | ADHD  ODD | Semi-structured child psychiatric interview (the Kiddie-SADS-PL) administered to a parent | baseline age 3-4 years | Child’s age, gender, SES, birth weight, and ethnicity, self-report of maternal and paternal ADHD symptoms, and maternal alcohol use during pregnancy | **OR (95% CI), p-value**  ADHD: 4 (1.36, 11.12), 0.012  ODD: 3.37 (0.22, 38.46), 0.34  **stronger association with maternal smoking compared to paternal smoking in ADHD* |
| Fergusson et al., 1998 | New Zealand | prospective longitudinal cohort study (the Christchurch Health and Development Study) | -/1022 | both | 16-18 years | Prospective self-report during each pregnancy trimesters | No smoking;  1-9 cigarettes/day; 10-19 cigarettes/day;  20+ cigarettes/day | CD | the Composite International Diagnostic Interview and the Self-Report Delinquency Inventory. DSM-IV criteria were used for constructing diagnosis | At birth, 4 months, annual assessment up to 16 years and 18 years | Maternal education, age, family SES, pregnancy planning, prenatal alcohol and illicit drug use, maternal child-rearing practices, parental separation, parental conflict, parental history of alcohol problems, criminal offending and illicit drug use | **OR (95% CI), SE, p-value** 1.27 (0.9, 1.78), 0.17, 0.172  **exposure treated in continuous scale* |
| Koshy et al., 2011 | UK | cross-sectional study (preselected areas of lower socio-economic status) | 29/1074 | both | 7.3 years | retrospective self-report (pregnancy trimester not specified) | No smoking (ref); Light: <10 cigarettes/day; Heavy: >10 cigarettes/day | ADHD | Parent report whether child has been diagnosed for ADHD by a doctor | NA | child obesity and overweight, doctor-diagnosed asthma, preterm birth, household member smoking during pregnancy, low birthweight | **OR (95% CI), p-value**  Total smoking: 3.19 (1.08, 9.49), 0.04  Light: 2.89 (0.38, 22.09) Heavy: 10.03 (1.62, 61.99), 0.013 |
| Pohlabeln et al., 2017 | Multiple countries (Belgium, Cyprus, Estonia, Hungary, Italy, Spain, Sweden) | prospective cohort study in general population (the European IDEFICS study – Identification and prevention of dietary- and lifestyle-induced health effects in children and infants) | 152/15,277 | both | 6.2 years | retrospective self-report (pregnancy trimester not specified) | Never (ref);  <1 a month;  Several occasions a week/daily | ADHD | Maternal report – “Has the child ever been diagnosed for ADHD? | 2 years (baseline age range 2-11.9 years) | child's sex, age, country, maternal age, SES and further for gestational hypertension, proteinuria, sugar in urine (glycosuria), and gestational diabetes, weight and height of the child at birth, potential health problems (respiratory adjustment disorders or infections), the duration of breastfeeding (exclusive and in combination with other types of feeding), and, in case of preterm birth, the number of weeks the child was born before the estimated date of birth. | **OR (95% CI), p-value**  Several occasions week/daily: 1.74 (1.13, 2.67), 0.02  **adjusted results reported only for heavier smoking* |
| Sagiv et al., 2013 | USA | prospective cohort study in general population (the New Bedford Cohort study) | 75/604 | both | 8 years | retrospective self-report (pregnancy trimester not specified) | No smoking (ref);  1-10 cigarettes/day >10 cigarettes/day | ADHD | Diagnosis were obtained from the paediatric medical records and parents were asked to report whether the child was regularly taking ADHD medications | 8 years | maternal age at offspring birth and education, paternal education, annual household income, marital status, prenatal substance use, maternal IQ, maternal depression (postnatal), HOME score, child age, gestational age, sex, ethnicity, breast feeding, school type, number of siblings | **RR (95% CI), p-value**  1-10 cigarettes:  0.9 (0.4, 1.8), 0.68  >10 cigarettes:  1.6 (0.8, 3.2), 0.20 |
| Schmitt et al., 2012 | Germany | cross-sectional study in general population (the German Health Interview and Examination Survey for Children and Adolescents (KIGGS) | 660/13,488 | both | 9.9 years | not specified | Binary (Yes/No) | ADHD | lifetime diagnosis of ADHD on the basis of medical or psychological examination as reported in standardized parental interviews conducted by trained interviewers | NA | child gender, age, SES (based on parental education, professional qualification, family income), maternal gestational diabetes, perinatal health problems, breastfeeding, atopic eczema | **OR (95% CI)**  1.48 (1.19, 1.84) |
| Sciberras et al., 2011 | Australia | population-based longitudinal study (the Longitudinal Study of Australian Children (LSAC) | 64/3781 | both | 6.8 years | retrospective self-report (pregnancy trimester not specified) | No smoking (ref); Occasionally; Most days | ADHD | Primary caregiver report – “Does the study child have ADHD?” | 2 years (baseline age 4-5 years) | maternal age, education, marital status, number of people in the household, child gender, household income, child's birth weight, maternal postnatal depression, intensive care at birth | **OR (95% CI), p-value** Occasionally:  0.62 (0.17, 2.26)  Most days:  3.31 (1.49, 7.39), 0.004 |
| Lehn et al., 2007 | the Netherlands | longitudinal twin study (the Netherlands Twin Registry) | 34/190 | both | 13.4 years | retrospective self-report (pregnancy trimester not specified) | Binary (Yes/No) | ADHD | Clinical Interview (DISC-IV Parent Version) | assessment at age 1, 2, 3, 5, 7, 10 and 12 years | Discordant and concordant pairs were matched on gender, zygosity, date of birth, maternal age, and parental SES. In wave II matching criteria was gender, date of birth, zygosity, handedness, and SES | **prenatal maternal smoking was more common in MZ pairs concordant for APs/ADHD than in the other groups, providing at least secondary evidence that smoking, in addition to a common genetic risk, leads to increased rates of ADHD in offspring* |
| Russell et al., 2015 | USA | cross-sectional study (household survey in general population) | -/5924 | both (reported separately for boys and girls) | 15.3 years | retrospective self-report (pregnancy trimester not specified) | More than 20 cigarettes per day; 11 to 20 cigarettes per day;  1 to 10 cigarettes per day;  Fewer than 1 cigarette per day; None (ref) | ODD | Structured diagnostic interview (CIDI - The World Health Organization’s Composite International Diagnostic Interview) | NA | offspring age | **OR (95% CI)**  Total sample: 1.06 (0.8, 1.3) Boys: 1.00 (0.7, 1.4)  Girls: 1.11 (0.8, 1.5)" |
| Froehlich et al., 2009 | USA | cross-sectional study (household survey in general population) | 215/2,588 | both | 8-15 years | retrospective self-report (pregnancy trimester not specified) | Binary (Yes/No) | ADHD | Diagnostic Interview Schedule for Children (DISC) and maternal report "Has a doctor or health professional ever told you that your child had attention deficit disorder" and ADHD medication treatment | NA | child age, ethnicity and gender, household income/poverty ratio, mother's age at child's birth, birth weight, NICU admission, postnatal secondhand smoke exposure, preschool attendance, health insurance status | **OR (95% CI), p-value** 2.4 (1.5-3.7), 0.001 |
| Knopik et al., 2005 | USA | longitudinal twin study (the Missouri Adolescent Female Twin study) | 128/1936 | female | 14.4 years (mean) | retrospective interview (assessed 1st pregnancy trimester and beyond 1st trimester) | First trimester: 1-10 cigarettes/day; 11-19 cigarettes/day; >20 cigarettes/day Beyond first trimester: 1-10 cigarettes/day; 11-19 cigarettes/day; >20 cigarettes/day | ADHD | maternal interview - the Diagnostic Interview for Children and Adolescents (DICA and the C-SSAGA (Semi-Structured Assessment of the Genetics of Alcoholism – Child Version) | recruitment over 2 years - 13, 15, 17 and 19 years | zygosity, prenatal and parental predictors - low birth weight, maternal alcohol abuse/dependence, paternal alcohol dependence, frequent heavy drinking during pregnancy | **OR (95%CI)** First trimester: 0.97 (0.5-1.86) 1-10 cigarettes/day: 1.05 (0.48-2.37) 11-19 cigarettes/day: 0.42 (0.11-1.63) >20 cigarettes/day: 1.40 (0.48-4.07)  Beyond first trimester: 1.50 (0.86-2.63) 1-10 cigarettes/day: 1.24 (0.61-2.52) 11-19 cigarettes/day: 1.83 (0.89=3.76) >20 cigarettes/day: 1.79 (0.79-4.07) |
| Knopik et al., 2006 | Australia | longitudinal twin study | 922 | both | 13-21 years | retrospective interview (assessed 1st pregnancy trimester and beyond 1st trimester) | Never smoked; Regular smoker, not during pregnancy;  1st trimester only Beyond 1st trimester, 1–15 cigarettes/day; Beyond 1st trimester, 16+ cigarettes/day | ADHD | maternal interview - the Diagnostic Interview for Children and Adolescents (DICA and the C-SSAGA (Semi-Structured Assessment of the Genetics of Alcoholism – Child Version) |  | child gender and age, paternal history of alcohol problems and paternal conduct disorder/antisociality history | **OR (95% CI), p-value** Never smoked (ref) Regular smoker, not during pregnancy: 0.72 (0.23-2.22) 1st trimester only:  1.88 (0.45-7.81) Beyond 1st trimester, 1–15 cigarettes/day:  0.54 (0.16-1.83) Beyond 1st trimester, 16+ cigarettes/day:  3.83 (1.09-13.45), <0.005 |
| Schwenke et al., 2018 | Germany | hospital based cohort study | 43/573 | both | 10 years | prospective data collection during pregnancy at the hospital | Binary (Yes/No) | ADHD | maternal report whether the child was taking medication for ADHD | 10 years | Apgar score | **OR (95% CI)** 2.63 (1.39, 4.97) |
| **Alcohol** |  |  |  |  |  |  |  |  |  |  |  |  |
| Fergusson et al., 1998 | New Zealand | prospective longitudinal cohort study (the Christchurch Health and Development Study) | -/1022 | both | 16-18 years | Prospective self-report during each pregnancy trimester | No drinking (ref);  1-3 drinks/week;  4-6 drinks/week;  7+ drinks/week | CD | the Composite International Diagnostic Interview and the Self-Report Delinquency Inventory. DSM-IV criteria were used for constructing diagnosis | At birth, 4 months, annual assessment up to 16 years and 18 years | Maternal education, age, family SES, pregnancy planning, prenatal smoking and illicit drug use, maternal child-rearing practices, parental separation, parental conflict, parental history of alcohol problems, criminal offending and illicit drug use | **OR (95% CI), SE, p-value** 1.32 (0.93, 1.88), 0.18, 0.126  **exposure treated in continuous scale* |
| Larkby et al., 2011 | USA | longitudinal birth cohort study | 67/487 | both | 16.8 years | Prospective self-report in each pregnancy trimester (focus on 1st and 3rd pregnancy trimester) | Alcohol consumption based on average daily volume (ADV)  No drinking (ref); Light (ADV <0.4); Moderate (0.4-0.89);  Heavy (>0.89) | CD | the Diagnostic Interview Schedule-IV (DIS-IV). | At delivery, 8 and 18 months, 3, 6, 10, 14, 16 and 22 years | prenatal exposure to marijuana, cocaine, and other illicit drugs, income, child's race, age and gender, parenting style, life events, home environment, family history of alcohol problems, and maternal lifetime psychopathology | **OR (95% CI)**  First trimester:  Heavy: 2.47 (1.3, 4.7)  **Results not reported for other categories and drinking in the 3rd trimester* |
| Pohlabeln et al., 2017 | Multiple countries (Belgium, Cyprus, Estonia, Hungary, Italy, Spain, Sweden) | prospective cohort study in general population (the European IDEFICS study – Identification and prevention of dietary- and lifestyle-induced health effects in children and infants) | 152/15,277 | both | 6.2 years | retrospective self-report (pregnancy trimester not specified) | Never (ref);  <1 a month;  Several occasions a month/week | ADHD | Maternal report – “Has the child ever been diagnosed for ADHD?” | 2 years (baseline age range 2-11.9 years) | child's sex, age, country, maternal age, SES and further for gestational hypertension, proteinuria, sugar in urine (glycosuria), and gestational diabetes, weight and height of the child at birth, potential health problems (respiratory adjustment disorders or infections), the duration of breastfeeding (exclusive and in combination with other types of feeding), and, in case of preterm birth, the number of weeks the child was born before the estimated date of birth. | **OR (95% CI), p-value**  Several occasions month/week:  0.76 (0.23, 2.48), 0.65  **adjusted results reported only for heavier drinking* |
| Sagiv et al., 2013 | USA | prospective cohort study in general population (the New Bedford Cohort study) | 75/604 | both | 8 years | prospective self-report 2 weeks after child's birth (pregnancy trimester not specified) | <1 serving/month (ref); 1-2 servings/month >2 servings/month | ADHD | Diagnosis were obtained from the paediatric medical records and parents were asked to report whether the child was regularly taking ADHD medications | 8 years | maternal age at offspring birth and education, paternal education, annual household income, marital status, prenatal substance use, maternal IQ, maternal depression (postnatal), HOME score, child age, gestational age, sex, ethnicity, breast feeding, school type, number of siblings | **RR (95% CI), p-value** 1-2 servings: 2.5 (0.8, 7.2), 0.1 >2 servings: 0.8 (0.3, 2.1), 0.71 |
| Schmitt et al., 2012 | Germany | cross-sectional study in general population  (the German Health Interview and Examination Survey for Children and Adolescents (KIGGS) | 660/13,488 | both | 9.9 years | not specified | Binary (Yes/No) | ADHD | lifetime diagnosis of ADHD on the basis of medical or psychological examination as reported in standardized parental interviews conducted by trained interviewers | NA | child gender, age, SES (based on parental education, professional qualification, family income), maternal gestational diabetes, perinatal health problems, breastfeeding, atopic eczema | **OR (95% CI)** 1.02 (0.79, 1.33) |
| Whitbeck et al., 2009 | USA | cohort study (lagged sequential) (population based on indigenous culture) | -/546 | both | 10-15 years | retrospective self-report (pregnancy trimester not specified) | Binary (Yes/No) | CD | diagnostic interview (the Diagnostic Interview Schedule for Children-Revised (DISC-R) with parents and children | 4 annual assessments (baseline age 10-12 years) | child age, gender, marital status, income, parenting behaviour (maternal warmth, support and maternal approval) | **OR, p-value** Binge drinking: 3.29, <0.01 |
| Russell et al., 2015 | USA | cross-sectional study (household survey in general population) | -/5924 | both  (reported separately for boys and girls) | 15.3 years | retrospective self-report (pregnancy trimester not specified) | Everyday;  3 to 5 times per week;  1 to 2 times per week;  1 to 3 times per month;  Less than once per month; Never (ref) | ODD | Structured diagnostic interview (CIDI - The World Health Organization’s Composite International Diagnostic Interview) | NA | child age | **OR (95% CI)** Total sample: 0.56 (0.3, 1.1) Boys: 0.56 (0.2, 1.5) Girls: 0.52 (0.2, 1.5) |
| Eilertsen et al., 2017 | Norway | prospective longitudinal birth cohort study  (MoBa) | 220/34,283 | both | 5 years | prospective self-report (assessed during the 1st pregnancy trimester) | continuous score of AUDIT scale | ADHD | Diagnosis of hyperkinetic disorder obtained from the National Patient Registry | 6, 18 months, 3, 5, 7, 8 and 13 years (ongoing study) | parental education and income, maternal smoking during pregnancy, children’s birth order and children’s gender | **OR (95% CI)** 0.97 (0.93, 1.01) |
| Knopik et al., 2005 | USA | longitudinal twin study (the Missouri Adolescent Female Twin study) | 128/1936 | female | 14.4 years (mean) | retrospective interview (assessed 1st pregnancy trimester and beyond 1st trimester) | 1-10 days;  11-35 days; >35 days Some heavy alcohol use; Frequent heavy alcohol use | ADHD | maternal interview - the Diagnostic Interview for Children and Adolescents (DICA and the C-SSAGA (Semi-Structured Assessment of the Genetics of Alcoholism – Child Version) | recruitment over 2 years - 13, 15, 17 and 19 years | zygosity, prenatal and parental predictors - low birth weight, maternal alcohol abuse/dependence, paternal alcohol dependence, frequent heavy drinking during pregnancy | **OR (95% CI)** 1-10 days: 1.11 (0.72-1.71) 11-35 days: 0.97 (0.26-3.64) >35 days: 3.31 (0.83-13.12) Some heavy alcohol use: 2.20 (0.95-5.09) Frequent heavy alcohol use: 4.64 (1.40-15.50) |
| Lees et al., 2020 | USA | longitudinal study  (Adolescent Brain Cognitive Development Study (ABCD)) | 9,719 ADHD cases: 1,870 CD cases: 271 ODD cases: 1,283 | both | 9.9 years | retrospective self-report (pregnancy trimester not specified) | Binary (Yes/No); abstinent  light reducers - 2.3 drinks/week for the first 7 weeks; light, stable - 1.1 drinks/week throughout pregnancy; heavier reducers - 5.3 drinks/week for the first 7 weeks | ADHD CD ODD | maternal report of the Schedule for Affective Disorders and Schizophrenia for School-Age Children (K-SADS), based on DSM-5 criteria | 1 year | Child: birth weight, prematurity, child gender and ethnicity, child age at assessment, school grades Mother: age at child's birth, history of maternal depression, other substance use during pregnancy, education | **Beta (95% CI), p-value** ADHD Binary:  0.09 (0.02, 0.15), 0.17 abstinent (ref) Light reducer:  0.03 (-0.06, 0.12), 0.71 Stable/light:  0.39 (0.01, 0.66), 0.15 Heavier reducer:  0.22 (0.11, 0.32), 0.04 CD Binary:  0.03 (-0.12, 0.18), 0.84 **no dose-dependent results were calculated as binary and continuous exposure results were not significant* ODD Binary:  0.16 (0.09, 0.23), 0.03 Light reducer:  0.11 (0.01, 0.20), 0.26 Stable/light:  -0.01 (-0.33, 0.32), 0.99 Heavier reducers:  0.26 (0.15, 0.37), 0.02 |
| Mitchell et al., 2020 | UK | cohort study (Millenium Cohort Study) | 167/13,004 | both | 7 years | maternal self-report 9 months after child's birth | No drinking (ref); Light: 3 to 7 units per week; Moderate: 8 to 14 units per week; Heavy: >14 units per week or per occasion | ADHD | Parent report whether child has been diagnosed for ADHD by a doctor or other healthcare professional | 9 months, 3, 5 and 7 years | child gender, gestational age at delivery, parity, maternal and paternal age, maternal smoking status, maternal pre-pregnancy BMI, household income, maternal education, ethnicity and marital status | **OR (95% CI)** Never drinkers (ref) Light: 0.48 (0.53, 1.22) Moderate: 0.83 (0.40, 1.74) Heavy: 1.27(0.54, 2.98) |
| Pagnin et al., 2019 | Brazil | prospective longitudinal study (Gest Alcohol) | 20/81 | both | 12 years | Prospective self-report during all the pregnancy trimesters | Binary: Alcohol use in all trimesters; Binge drinking | ADHD | maternal report of the Schedule for Affective Disorders and Schizophrenia for School-Age Children - present and lifetime version (K-SADS) | 12 years | gender, current maternal mental disorders | **OR (95% CI)** Alcohol use in all trimesters:  5.11 (1.42, 18.38) Binge drinking:  4.72 (1.04, 21.47) |
| Weile et al., 2020 | Denmark | prospective cohort study (Aarhus Birth cohort) | 48,072 Binge drinking: 1,159 cases Weekly alcohol use: 1,058 cases | both | 12 years | prospective data collection during pregnancy (median 11 weeks) | Binge drinking:  0 episodes; 1 episode; 2 episodes; >3 episodes Average weekly alcohol intake: 0 drinks/week; <1 drink/week; 1 drink/week 2 drinks/week >3 drinks/week | ADHD | diagnosis obtained from Danish health registries / Danish National Patient Register for Psychiatry and Danish National Patient Register | up to 19.8 years | maternal age at birth, education, pre-pregnancy BMI, chronic disease, smoking in pregnancy, parity, birth year, alcohol use before pregnancy, child gender and maternal mental health diagnoses before the time of birth | Hazard ratio (HR) (95% CI) Binge drinking  0 episodes (ref) 1 episode: 0.91 (0.76,1.08) 2 episodes: 0.73 (0.56, 0.96) >3 episodes:  0.77 (0.57, 1.06) Average weekly alcohol intake 0 drinks/week (ref) <1 drink/week:  0.87 (0.74, 1.03) 1 drink/week:  0.63 (0.40, 0.98) 2 drinks/week:  1.30 (0.89, 1.92) >3 drinks/week:  0.78 (0.38, 1.59) |
| **Caffeine** |  |  |  |  |  |  |  |  |  |  |  |  |
| Del-Ponte et al., 2016 | Brazil | population based longitudinal cohort study (Pelotas birth cohort study) | 142/3485 | both  (reported separately for boys and girls) | 11 years | Prospective interview after child’s birth (assessed each trimester and throughout pregnancy) | <100 mg/day (ref); 100-299 mg/day; >300 mg/day | ADHD | The Development and Well-Being Assessment Scale (DAWBA) reported by mothers and diagnosis by a child psychiatrist | 3, 12, 24 and 48 months, 6 and 11 years (ongoing study) | National Economic Index mother’s and father’s education levels, evaluated as years of study; maternal age, mother living with or without partner; number of cigarettes smoked per day by the mother during pregnancy; number of cigarettes smoked per day by the father in the mother’s presence during pregnancy; alcohol consumption by the mother during pregnancy; number of antenatal care consultations; mood symptoms during pregnancy; maternal nutritional state before pregnancy, evaluated according to the body mass index (BMI); the child gestational age at birth; birth weight and sex | **OR (95% CI)** 1st trimester total sample  100-299 mg/day:  1.04 (0.62, 1.73) >300 mg/day:  0.93 (0.55, 1.60) Boys 100-299 mg/day:  1.06 (0.57, 1.98) >300 mg/day:  1.06 (0.57, 1.96) Girls 100-299 mg/day:  1.13 (0.44, 2.90) >300 mg/day:  0.68 (0.21, 2.17) 2nd trimester total sample  100-299 mg/day:  1.03 (0.61, 1.74) >300 mg/day:  0.95 (0.55, 1.63) Boys 100-299 mg/day:  1.03 (0.55, 1.94) >300 mg/day:  1.09 (0.58, 2.03) Girls 100-299 mg/day:  1.24 (0.48, 3.22) >300 mg/day:  0.75 (0.24, 2.38) 3rd trimester total sample  100-299 mg/day:  0.96 (0.55, 1.68) >300 mg/day:  1.05 (0.61, 1.81) Boys 100-299 mg/day:  0.82 (0.40, 1.68) >300 mg/day:  1.07 (0.57, 2.02) Girls 100-299 mg/day:  1.68 (0.64, 4.40) >300 mg/day:  1.22 (0.41, 3.60) Entire pregnancy total sample  100-299 mg/day:  1.12 (0.68, 1.84) >300 mg/day:  0.90 (0.51, 1.59) Boys 100-299 mg/day:  1.05 (0.57, 1.92) >300 mg/day:  1.01 (0.52, 1.95) Girls 100-299 mg/day:  1.46 (0.58, 3.68) >300 mg/day:  0.82 (0.25, 2.65) |
| Linnet et al., 2008 | Denmark | population-based longitudinal cohort study (the Aarhus Birth Cohort) | 88/24,068 | both | 7 years | prospective self-report (assessed during the 2nd pregnancy trimester) | No coffee (ref): 1-3 cups; 4-9 cups; 10+ cups | ADHD | Diagnosis was obtained from the Danish Psychiatric Case Register | up to age 12 years | prenatal smoking and alcohol use, maternal age, gender of the child, parental years of schooling after basic school, employment status, cohabitant status and parental and siblings’ psychiatric hospitalisations or contacts in outpatient clinics | **OR (95% CI)** 1-3 cups: 0.9 (0.5, 1.6) 4-9 cups: 1.3 (0.7, 2.3) 10+ cups: 2.3 (0.9, 5.9) |
| Russell et al., 2015 | USA | cross-sectional study (household survey in general population) | -/5924 | both  (reported separately for boys and girls) | 15.3 years | retrospective self-report (pregnancy trimester not specified) | None to less than one cup per day; One or more cups per day | ODD | Structured diagnostic interview (CIDI - The World Health Organization’s Composite International Diagnostic Interview) | NA | child age | **OR (95% CI)** Total sample: 0.86 (0.7, 1.0) Boys: 0.98 (0.8, 1.3) Girls: 0.75 (0.6, 1.0), p=<0.05*  **coding unclear (authors are reporting increasing effect on ODD of caffeine use in girls)* |

**studies included in meta-analysis*

### Supplementary Table S6. Study characteristics and results of case-control studies

| **Author/Year** | **Country** | **Study sample** | **Cases/controls** | **Gender** | **Offspring age at assessment** | **Exposure assessment** | **Classification of exposure** | **Outcome** | **Outcome assessment** | **Follow up time** | **Confounders** | **Results (as reported in the study)** |
| --- | --- | --- | --- | --- | --- | --- | --- | --- | --- | --- | --- | --- |
| **Smoking** |  |  |  |  |  |  |  |  |  |  |  |  |
| Biederman et al., 2009 | USA | longitudinal case-control family study (clinical population) | 291/536 | both | 13.2 years | retrospective interview (pregnancy trimester not specified) | Binary (Yes/No) | ADHD  CD | Clinical interview (K-SADS-E and SCID) with mothers and offspring, diagnosis confirmed by a psychiatrist | Assessments at baseline, 4 and 10 years for boys and in 5 years in girls. Age range at baseline 5 – 37 years (mean 13.4 years) | maternal age at offspring birth, social class, offspring age at baseline, offspring gender, parental lifetime history of ADHD, parental lifetime history of CD, prenatal exposure to maternal alcohol or illicit drugs, study of origin (Boys ADHD, Girls ADHD), referral status, number of assessments (ADHD analyses were adjusted for CD and vice versa) | **OR (95% CI), p-value**  ADHD: 2.5 (1.39, 4.51), 0.002  CD: 3.3 (1.23, 8.88)  **the association with CD was only among siblings from control families. No association was observed among siblings of ADHD families* |
| Gard et al., 2016 | USA | longitudinal case-control (clinical and community sample) | 140/88 | only girls | 20 years | retrospective self-report (pregnancy trimester not specified) | Binary (Yes/No) | ADHD hyperactive-impulsive and inattention subtype | Diagnostic interview (DISC-IV) with mothers as well as maternal and self-report on ADHD symptom domains (diagnosis based on DSM-IV criteria) | 2 waves – 5 (mean age 14 years) and 10 years (mean age 20 years) | SES, maternal education, income, maternal ADHD, participant ODD symptoms and substance use  **cases and controls matched by age and ethnicity* | **Beta, p-value**  Maternal report:  HYP: 0.16, 0.03  INA: 0.10  Self-report:  HYP: 0.13, 0.08  INA: 0.09 |
| Gustafsson and Kallen, 2010 | Sweden | Population-based case-control registry study | 229/32,012 | Both | 8-12 years | prospective self-report during antenatal visit (pregnancy trimester not specified) | No smoking;  <10 cigarettes/day; >10 cigarettes/day | ADHD | Diagnosis derived from the hospital database (the department of child and adolescent psychiatry in Malmo) | from 5 up to 17 years | birth year, maternal age, birthplace, preterm birth, Apgar score, gestational age, child gender | **OR (95% CI), p-value**  1.35 (1.14, 1.6) |
| Joelsson et al., 2016* | Finland | population-based nested case-control registry study | 3136/50,550 | Both | not specified | Prospective self-report to maternity clinic during 2nd pregnancy trimester | Non-smokers; Smoking only 1st trimester;  Smoking after 1st trimester (analyses with binary exposure) | ADHD (with and without comorbid CD/ODD cases) | the Finnish Hospital Discharge Register (FHDR) | not specified | maternal and paternal psychiatric history, maternal history of any substance use, maternal and paternal age at birth of offspring, maternal and paternal immigrant status, maternal socioeconomic status (SES), birth weight for gestational age, Apgar scores at 1 min, number of previous births and gestational age  **cases and controls matched by date of birth, sex and residence in Finland* | **OR (95% CI)**  1.59 (1.43, 1.77) |
| Linnet et al., 2005* | Denmark | nested case-control registry study | 170/3935 | both | 5.5 years | Prospective self-report collected by midwives during antenatal visit (pregnancy trimester not specified) | Binary (Yes/No) | Hyperkinetic disorder | Diagnosis were obtained from the Danish Psychiatric Central Register | up to 8 years | maternal, paternal, and siblings’ psychiatric hospitalizations and outpatient contacts, socioeconomic factors, and maternal age  **cases and control subjects were matched for age, gender, and calendar time* | **Relative Risk (RR) (95% CI)** 1.9 (1.3, 2.8) |
| Milberger et al., 1996 | USA | case-control family study (clinical population) | 140/260 | only boys | 6-17 years | retrospective interview (pregnancy trimester not specified) | Binary (Yes/No) | ADHD | Structured interviews with mothers and offspring’s using the childhood version of the Schedule for Affective Disorders and Schizophrenia for School-Age Children – Epidemiologic Version (K-SADS-E). Diagnosis were confirmed by the psychiatrist | not specified | socioeconomic status, parental ADHD status, and parental IQ  **cases and controls matched by ethnicity, gender and age* | **OR (SE), (95% CI), p-value** 2.7 (1.31), (1.1, 7), 0.04 |
| Milberger et al., 1998 | USA | case-control family study (clinical population) siblings of ADHD and non-ADHD families | 174/300 | both | 13.5 years | retrospective interview (pregnancy trimester not specified) | Binary (Yes/No) | ADHD | Structured interviews with mothers and offspring’s using the childhood version of the Schedule for Affective Disorders and Schizophrenia for School-Age Children – Epidemiologic Version (K-SADS-E). Diagnosis were confirmed by the psychiatrist | not specified | socioeconomic status, parental ADHD status, and parental IQ  **cases and controls matched by ethnicity, gender and age* | **OR (SE), (95% CI), p-value** 4.4 (2.8), (1.2, 15.5), 0.02 |
| Schmitz et al., 2006 | Brazil | case-control | 100/200 | both | 11.8 years | retrospective interview (pregnancy trimester not specified) | No smoking (ref);  1-9 cigarettes/day; >10 cigarettes/day | ADHD inattention subtype | 3 stage process: The first stage was a semi-structured interview (Schedule for Affective Disorders and Schizophrenia for School-Age Children, Epidemiological Version [K-SADS-E] administered to the parents. In stage 2, each diagnosis derived from the K-SADS-E was discussed in a clinical committee chaired by an experienced child and adolescent psychiatrist. For the third stage, a clinical evaluation of ADHD-I and comorbid conditions using DSM-IV criteria was conducted by a child and adolescent psychiatrist who previously received the results of the K-SADS-E and conducted interviews with the parents and the child or adolescent. | NA | maternal age, SES, maternal ADHD, child ODD, alcohol use in pregnancy, birth weight  **cases and controls matched by gender and age* | **OR (95% CI), p-value**  No smoking (ref)  1-9 cigarettes:  1.09 (0.32, 3.66), 0.89  >10 cigarettes:  3.44 (1.17, 10.06), 0.02 |
| Silva et al., 2014 | Australia | population-based case-control record linkage study | 12,911/43,062 | Both (reported separately for boys and girls) | <25 years | Prospective self-report collected by midwives during antenatal visit (pregnancy trimester not specified) | Binary (Yes/No) | ADHD | stimulant medication for ADHD identified through the Monitoring of Drugs of Dependence System (MODDS). Controls were selected from the Midwives Notification System (MNS) and matched by birth year, gender and SES | up to 25 years | marital status, parity, pregnancy complications, onset of labour, complications of labour, type of delivery, child gestational age, birth weight *cases and controls matched by year of birth, gender and SES | **OR (95% CI)**  Boys: 1.86 (1.53, 2.27)  Girls: 1.67 (1.07, 2.61)" |
| Todd et al., 2007 | USA | case-control (population-based twin study, MOTWIN) | 198/1518 | both | 13 years | retrospective self-report (assessed each pregnancy trimester) | Binary (Yes/No) | ADHD combined and inattentive subtype | Parents interview using the Missouri Assessment of Genetics Interview for Children (MAGIC) to obtain DSM-IV diagnoses and individual symptom information. | NA | child gender, negative expressed emotion in the family and DSM-IV diagnoses of oppositional defiant disorder (ODD) and conduct disorder (CD) | **OR (95% CI)**  ADHD_combined_: 3.9 (1.2, 13.1)  **the association was observed between prenatal smoking and child genotype at the rs1044396 C allele (CHRNA4 gene) **Association was also observed between prenatal smoking and two genes (CHRNA4 and DAT1) - ADHD_comb_ OR=6 (1.2, 28.9)* |
| Yoshimasu et al., 2009 | Japan | case-control (hospital-based population) | 90/360 | both | 10 years | retrospective interview (pregnancy trimester not specified) | Lifetime non-smokers (ref); Former smokers; Stopped smoking when aware of pregnancy; Continued smoking | ADHD | experienced psychiatrists or paediatricians diagnosed ADHD according to the diagnostic criteria of DSM-IV, with full consideration of both parents’ and teachers’ evaluations. | NA | children’s gender, family income, maternal drinking during pregnancy, pregnancy-induced hypertension, birth weight, and children’s iron intake, maternal tendency of ADHD, parental history of mental disorders and maternal mental stress during pregnancy  **cases and controls matched by age* | **OR (95% CI)**  Lifetime non-smokers (ref) Former smokers:  0.8 (0.3, 2.5)  Stopped smoking:  1.3 (0.5, 3.6)  Continued smoking:  1.3 (0.5, 3.6)" |
| Altink et al., 2009 | the Netherlands (the International Multi-centre ADHD Gene project (IMAGE)) | case-control family study | 79/184 | both | 12 years | retrospective self-report (assessed each pregnancy trimester) | Binary (Yes/No) | ADHD | All probands were included after completing clinical evaluations by a paediatrician or child psychiatrist prior to the study. The clinical diagnosis of the ADHD probands and siblings was verified with the Parental Account of Childhood Symptoms (PACS) by a trained interviewer | NA | Child age, gender, IQ, birth weight, oppositional and anxious-shy symptoms of a child, total maternal or paternal ADHD symptoms, maternal age and socio-economic status | **OR (95% CI), p-value** *association between maternal prenatal smoking and ADHD status  3.29 (1.48, 7.30), 0.003  This relationship was partly mediated by the performance on attentional control:  OR=2.42, 95%CI 1.04-5.61), p=<0.001  **no effect was found between prenatal smoking and child genotype on attentional control in children with ADHD*  **paternal risk genes (DRD4, DAT1) mediated paternal smoking and effect on attentional control in children with ADHD* |
| Altink et al., 2008 | Multiple countries (Belgium, Germany, Ireland, Spain, Switzerland, the Netherlands and UK) (IMAGE study) | case-control family study | 539/946 | both | 11 years | retrospective self-report (assessed each pregnancy trimester) | Binary (Yes/No) | ADHD | All probands were included after completing clinical evaluations by a paediatrician or child psychiatrist prior to the study. The clinical diagnosis of the ADHD probands and siblings was verified with the Parental Account of Childhood Symptoms (PACS) by a trained interviewer | NA | child gender, age and birth weight | **OR (95% CI), p-value** *association between prenatal smoking and ADHD status:  1.76 (1.09-2.85), 0.021  **no interaction effect was found between DRD4 gene and prenatal smoking 1.86 (0.69, 4.98)* |
| Neuman et al., 2007 | USA | case-control (population-based twin study) (MOTWIN sample) | 140/832 | both | 13 years | retrospective interview (pregnancy trimester not specified) | Binary (Yes/No) | ADHD combined and inattentive subtype | 1st stage - screening interview 2nd stage - diagnostic interview completed by parents (the Missouri Assessment of Genetics Interview for Children (MAGIC), a modified version of the Diagnostic Interview for Children and Adolescents) | NA | Child’s gender, negative home environment, ODD and CD diagnosis | **OR (95% CI)**  Any ADHD: 1.58 (1.03, 2.43) ADHD combined:  1.91 (0.97, 3.76)  ADHD inattention:  1.52 (0.89, 2.58)  **The odds for a diagnosis of DSM-IV ADHD was 1.8 times greater in twins whose genotype at the DAT3= VNTR contained the 440 allele and whose mother smoked during pregnancy than for twins who had neither risk factor. Similarly, the risk for a diagnosis of DSM-IV ADHD was significantly elevated in twins with prenatal smoke exposure and the DRD4 seven-repeat allele. There were no significant interactions for the DSM-IV ADHD phenotype between prenatal smoking and the DAT1 480 allele* |
| Arnold et al., 2005 | USA | case-control (the Multimodal Treatment Study of Children with ADHD) | 164/461 | both | 7-9.9 years | retrospective self-report (pregnancy trimester not specified) | Binary (Yes/No) | ADHD | diagnosed prior to the study by the clinician | 14 months | Child sex, mother’s educational level, public assistance, single parent status, prenatal drinking, family history of ADHD and CD | **p-value**  *prenatal smoking predicted ADHD status ADHD with comorbid CD/ODD: 0.024 ADHD without comorbiditeis: 0.096  **No moderating effect was found between prenatal smoke exposure and ADHD treatment outcome* |
| Biederman et al., 2017 | USA | case-control (family study clinical population) | 496  267 ADHD cases  115 CD cases 211 ODD cases" | both | 21 years | retrospective interview (pregnancy trimester not specified) | Binary (Yes/No) | CD  ODD | The assessment had 3 stages: referral; telephone questionnaire with mother; diagnostic interview (DISC) | 10 years for boys and 11 years for girls | SES, child ADHD **cases and controls matched by gender and age* | **OR (95% CI), p-value**  CD: 1.51 (0.66, 1.96), 0.13 ODD: 1.14 (0.66, 1.96), 0.63 |
| Ketzer et al., 2012 | Brazil | case-control | 124/248 | both | 11.8 years | retrospective self-report (pregnancy trimester not specified) | Binary (Yes/No) | ADHD inattention subtype | 3 stage assessment: 1st stage – a semi-structured interview (Schedule for Affective Disorders and Schizophrenia for School-Age Children) modified to assess DSM-IV criteria and administered to the parents by trained research assistants 2nd stage - each diagnosis derived from the K-SADS-E was discussed in a clinical committee chaired by an experienced child and adolescent psychiatrist 3rd stage - a clinical evaluation of ADHD-I and comorbid conditions were performed according to DSM-IV criteria by a child and adolescent psychiatrist who previously had access to K-SADS-E results. | NA | generalised anxiety disorder, ODD, agoraphobia, maternal ADHD. *Cases and controls were matched by age and gender | **OR (95% CI), SE, p-value**  1 (0.9, 1.1), 0.03, 0.2 " |
| Mick et al., 2002 | USA | case-control family study (clinical population) | 280/522 | both | 11 years | retrospective interview (pregnancy trimester not specified) | Binary (Yes/No) | ADHD | structured diagnostic interview (the Schedule for Affective Disorders and Schizophrenia for School-Age Children-Epidemiologic version (K-SADS-E). All assessments were made by raters who were blind to the child’s diagnosis (ADHD or non-ADHD control) and ascertainment site | NA | maternal age at child’s birth, indicators of social adversity (low social class, large family size, severe marital discord), parental history of ADHD, parental history of CD/ASPD, and comorbid CD in cases and controls *cases and controls matched by gender and age | **OR (95% CI), p-value**  2.1 (1.1, 4.1), 0.02" |
| Pineda et al., 2007 | Columbia | case-control | 200/486 | both | 8.3 years | retrospective self-report (pregnancy trimester not specified) | No smoking (ref); Low: 1-3 cigarettes/day  High: 4+ cigarettes/da | ADHD | psychiatric Diagnostic Interview for Children and Adolescents—parent revised Spanish version (DICA-PR). DSM-IV-ADHD-symptoms questionnaire, Behavioural Assessment System for Children (BASC) - parent and teacher report. All psychiatric, medical, neurological, neuropsychological, and psychological records of the ADHD children were reviewed by neurologists and neuro- psychologists. Parents provided information about the current clinical condition of their children and filled out the ADHD retrospective structured risk factor survey. | NA | gender and school grades *cases and controls matched by age, SES and school level | **OR (95% CI), p-value**  High: 8.9 (1.0, 78.4), <0.05 |
| Motlagh et al., 2010 | USA | case-control | 52/117 | both | 11.8 years | retrospective interview (pregnancy trimester not specified) | Binary (>10 cigarettes/day/No) | ADHD | Semi-structured interview using the Schedule for Affective Disorders and Schizophrenia for School-Age Children-Present and Lifetime Version (SADS-PLV) and a ‘‘best estimate’’ consensus procedure in which expert clinicians considered all available clinical and diagnostic information | NA | gender, severe psychosocial stress and limited coping abilities during pregnancy, more than one pregnancy complication, and antibiotic use *cases and controls matched by age and ZIP code | **OR (95% CI), p-value**  18.6 (2.1, 164.4), 0.008" |
| Wiggs et al., 2016 | USA | case-control family study | 251/464 | both | 11 years | retrospective self-report (pregnancy trimester not specified) | Binary (Yes/No) | ADHD (hyperactivity-impulsivity and inattention)  CD  ODD | Schedule for Affective Disorders and Schizophrenia-E for each child with a trained master’s level clinical interviewer. Clinical data were reviewed, and a best estimate diagnostic procedure was implemented by a board-certified child psychiatrist and a licensed child clinical psychologist | NA | child sex, age, ethnicity, income, parental ADHD symptoms and externalizing disorder status, and maternal and paternal age at birth | **Beta(β), 95% CI, p-value** Direct effect of prenatal tobacco exposure  INA: 0.05 (-0.01, 0.11), 0.19 HYP: 0.03 (-0.05, 0.11), 0.56 CD: 0.02 (-0.08, 0.12), 0.76 ODD: 0.04 (-0.06, 0.14), 0.19  **Indirect effect of prenatal tobacco exposure via neuropsychological functioning*  *INA: 0.08 (0.02, 0.14), 0.02 specifically via memory span β=.03, (0.005, .06], p=. 039*  **indirect effects were observed also for CD*  *β=.04, (0.005, .07], p=.060 and ODD β=.04, (0.002, .07], p=.08* |
| Oerlemans et al., 2016 | the Netherlands (the International Multi-centre ADHD Gene project (IMAGE)) | case-control family study | 201/768 | both | 11.2 years | retrospective self-report (pregnancy trimester not specified) | Binary (Yes/No) | ADHD | Conners rating scale; Parental Account of Childhood Symptoms ADHD subversion and diagnostic interview | NA | Family size, parity, child gender | **OR (95% CI), p-value**  2.12 (1.52-4.48), 0.005  **single and multiple ADHD incidence stratification showed that prenatal smoking was a shared familial risk between affected and unaffected ADHD children* |
| Wang et al., 2019 | China | case-control | 168/233 | both | 8.5 years | retrospective self-report  (pregnancy trimester not specified) | Binary (Yes/No) | ADHD combined, hyperactive and inattentive subtype | clinical interview based on the DSM-IV ADHD Rating scale and conducted by a psychiatrist | NA | age, gender,  birth weight, pregnancy age of mother, preterm birth, family history of neural system disease, blood lead concentration and postnatal tobacco smoke exposure | **OR (95% CI), p-value** ADHD:  1.75 (1.07, 2.94), <0.05 HYP: 2.34 (1.23, 4.47), <0.01 INA: 1.71 (1.01, 2.90), <0.05  **interaction effect was observed with smoking and genes (ADRA2A rs553668 AG+GG polymorphisms (OR=3.54; 95%CI 1.73, 7.25). In DRD2 rs1124491 AA (OR=5.34; 95%CI 1.64-4.46) and AG+GG (OR=6.04; 95%CI 2.07, 7.65) polymorphisms. As well as SLC6A4 rs6354 TT (OR=2.13; 95%CI 1.20-3.78) and TG+GG (OR=1.79; 95%CI 0.75-4.28) polymorphisms)* |
| Sourander et al., 2019 | Finland | nested case-control study | 1,079/1,079 | both | 7.3 years | prospective data collection during the 1st and 2nd pregnancy trimester | cotinine measure: continuous low (<20ng/mL) moderate: (20-50ng/mL) heavy: (>50ng/mL) | ADHD | The Finnish Medical Birth Register | NA | number of previous births, maternal SES, maternal psychiatric history, maternal history of ADHD diagnosis and substance use disorder, paternal history of ADHD diagnosis, gestational age of the child, birth weight for gestational age, maternal and paternal age **cases and controls were matched on date of birth, sex and place of birth* | **OR (95% CI), p-value** continuous:  1.09 (1.06, 1.13), <0.001 low (ref) moderate:  1.27 (0.84, 1.92), 0.25 heavy:  2.21 (1.64, 2.99), <0.001 |
| **Alcohol** |  |  |  |  |  |  |  |  |  |  |  |  |
| Ketzer et al., 2012 | Brazil | case-control | 124/248 | both | 11.8 years | retrospective self-report (pregnancy trimester not specified) | Binary (Yes/No) | ADHD inattention subtype | 3 stage assessment: 1st stage – a semi-structured interview (Schedule for Affective Disorders and Schizophrenia for School-Age Children) modified to assess DSM-IV criteria and administered to the parents by trained research assistants 2nd stage - each diagnosis derived from the K-SADS-E was discussed in a clinical committee chaired by an experienced child and adolescent psychiatrist 3rd stage - a clinical evaluation of ADHD-I and comorbid conditions were performed according to DSM-IV criteria by a child and adolescent psychiatrist who previously had access to K-SADS-E results. | NA | social phobia, ODD, maternal ADHD, IQ, tobacco use in pregnancy  **cases and controls matched by age and gender"* | **OR (95% CI), SE, p-value**  3.2 (0.8, 12.6), 0.7, 0.1 |
| Mick et al., 2002 | USA | case-control family study (clinical population) | 280/522 | both | 11 years | retrospective interview (pregnancy trimester not specified) | Binary (Yes/No) | ADHD | structured diagnostic interview (the Schedule for Affective Disorders and Schizophrenia for School-Age Children-Epidemiologic version (K-SADS-E). All assessments were made by raters who were blind to the child’s diagnosis (ADHD or non-ADHD control) and ascertainment site | NA | maternal age at child’s birth, indicators of social adversity (low social class, large family size, severe marital discord), parental history of ADHD, parental history of CD/ASPD, and comorbid CD in cases and controls | **OR (95% CI), p-value**  2.5 (1.1, 5.5), 0.03 |
| Pineda et al., 2007 | Columbia | case-control | 200/486 | both | 8.3 years | retrospective self-report (pregnancy trimester not specified) | No drinking (ref); Low: <10 drinks/week  High: Drunkenness during the first 2 months | ADHD | psychiatric Diagnostic Interview for Children and Adolescents—parent revised Spanish version (DICA-PR). DSM-IV-ADHD-symptoms questionnaire, Behavioural Assessment System for Children (BASC) - parent and teacher report. All psychiatric, medical, neurological, neuropsychological, and psychological records of the ADHD children were reviewed by neurologists and neuro- psychologists. Parents provided information about the current clinical condition of their children and filled out the ADHD retrospective structured risk factor survey. | NA | gender and school grades  **cases and controls matched by age, SES and school level.* | **OR (95% CI), p-value**  High: 11.7 (1.5, 94.1), 0.02 |
| Kim et al., 2009 | Korea | case-control | 100/2319 | both | 10.5 years | retrospective interview (pregnancy trimester not specified) | Binary (Yes/No) | ADHD | Psychiatric disorders were assessed, according to the Diagnostic and Statistical Manual of Mental Disorders 4th version (DSM-IV), with the Korean version of the DISC-IV and with parental interview | NA | age, gender, SES (by income) | **OR (95% CI)**  3.31 (1.59, 6.91) |
| **Caffeine** |  |  |  |  |  |  |  |  |  |  |  |  |
| Kim et al., 2009 | Korea | case-control | 100/2319 | both | 10.5 years | retrospective interview (pregnancy trimester not specified) | Binary (Yes/No) | ADHD | Psychiatric disorders were assessed, according to the Diagnostic and Statistical Manual of Mental Disorders 4th version (DSM-IV), with the Korean version of the DISC-IV and with parental interview | NA | age, gender, SES (by income) | **OR (95% CI)**  1.28 (0.81, 2.02) |

**Studies included in meta-analysis*

### Supplementary Table S7. Confounders included in the cohort, longitudinal and cross-sectional studies

| **Number of studies adjusted for this confounder** | | | |
| --- | --- | --- | --- |
| **Confounder** | **Smoking**  (29 studies) | **Alcohol**  (13 studies) | **Caffeine**  (3 studies) |
| Offspring gender | 21 (72%) | 11 (85%) | 2 (67%) |
| Offspring age | 18 (62%) | 7 (54%) | 1 |
| Offspring ethnicity | 9 (31%) | 5 (38%) | - |
| Offspring comorbid externalising disorders | - | - | - |
| Parity and/or number of siblings | 9 (31%) | 4 (31%) |  |
| Maternal age at offspring birth | 17 (59%) | 6 (46%) | 2 (67%) |
| Parental socio-economic characteristics (social class, education, income, marital status) | 21 (72%) | 10 (77%) | 2 (67%) |
| Parenting behaviour and/or home environment | 5 (17%) | 4 (31%) | - |
| Parental externalising disorder symptoms (ADHD, antisocial personality) | 3 (10%) | - | - |
| Other parental psychopathology and substance use disorders | 13 (45%) | 6 (46%) | 1 |
| Maternal mental health during pregnancy | 2 (7%) | 1 (8%) | 1 |
| Maternal other substance use during pregnancy | 10 (34%) | 7 (54%) | 2 (67%) |
| Partner’s or household member substance use during pregnancy | 1 (3%) | - | - |
| Perinatal factors (birth weight, gestational age, birth complications) | 15 (52%) | 6 (46%) | 1 |

### Supplementary Table S8. Confounders included in the case-control studies

| **Number of studies adjusted for this confounder** | | | |
| --- | --- | --- | --- |
| **Confounder** | **Smoking**  (24 studies) | **Alcohol**  (4 studies) | **Caffeine**  (1 study) |
| Offspring gender | 22 (92%) | 3 (75%) | 1 |
| Offspring age | 18 (75%) | 3 (75%) | 1 |
| Offspring ethnicity | 5 (21%) | - | - |
| Offspring comorbid externalising disorders | 9 (38%) | 2 (50%) |  |
| Parity and/or number of siblings | 4 (17%) | - | - |
| Maternal age at offspring birth | 10 (42%) | 1 |  |
| Parental socio-economic characteristics (social class, education, income, marital status) | 16 (67%) | 4 (100%) | 1 |
| Parenting behaviour and/or home environment | 2 (8%) | - | - |
| Parental externalising disorder symptoms (ADHD, antisocial personality) | 11 (46%) | 2 (50%) | - |
| Other parental psychopathology, and substance use disorders | 5 (21%) | - | - |
| Maternal mental health during pregnancy | 2 (8%) | - | - |
| Maternal other substance use during pregnancy | 5 (21%) | 1 |  |
| Partner’s substance use during pregnancy | - | - | - |
| Perinatal factors (birth weight, gestational age, birth and pregnancy complications) | 10 (42%) | - | - |
